# Echo chambers as early warning signals of widespread vaccine refusal in social-epidemiological networks

**DOI:** 10.1101/2020.10.17.20214312

**Authors:** Brendon Phillips, Chris T. Bauch

## Abstract

Sudden shifts in population health and vaccination rates occur as the dynamics of some epidemiological models go through a critical point; literature shows that this is sometimes foreshadowed by early warning signals (EWS). We investigate different structural measures of a network as candidate EWS of infectious disease outbreaks and changes in popular vaccine sentiment. We construct a multiplex disease model coupling infectious disease spread and social contact dynamics. We find that the number and mean size of echo chambers predict transitions in the infection dynamics, as do opinion-based communities. Graph modularity also gives early warnings, though the clustering coefficient shows no significant pre-outbreak changes. Change point tests applied to the EWS show decreasing efficacy as social norms strengthen. Therefore, many measures of social network connectivity can predict approaching critical changes in vaccine uptake and aggregate health, thereby providing valuable tools for improving public health.

## 1. Introduction

Low vaccine rates stemming from vaccine refusal result in outbreaks of vaccine-preventable diseases in some populations [1], and the high costs of intervention and treatment incurred by public health systems [2] motivate us to find tools warning of epidemics. The connection between social network activity and health issues in populations has long been exploited by researchers [3], especially relating to disease spread [4], with the assertion that firm understanding of social network structure is important to the implementation of effective policy interventions [5]. For example, vaccination decisions sometimes depend on communication and information diffusion in media [6]; as such, patterns of communication in social networks yield warning signals such as increased spatial autocorrelation [7].

Through assortative mixing, densely connected groups of members are formed in a social network with sparse connectivity between said groups; these groups are called *communities*, and greatly influence any dynamic process on the network [8]. Specifically, studying the formation and growth of communities on social networks allows for the discovery of non-obvious interrelationships [9]. Strongly connected components on social networks have been used as a proxy for community structure [10] as well as other structures and definitions [11].

Communities where every member shares the same opinion are called *opinion clusters* [8] or *opinion based communities* and are a fixture of online social networks, a direct result of assortative mixing [12]. Different methods of propagation of opinion have been studied in the literature: for example, neighbourhood sampling [13], summation and averaging [14], estimation [15] and population-level interaction [16]. In communication models where sentiment change is driven primarily by exposure to news sources and contrasting views from neighbours, communities can support the reinforcement of sentiments already held [17].

Echo chambers, described as well connected groups of people promoting and reinforcing the same bias [18], have recently come under media scrutiny since these groups can facilitate vaccine scares [18], support political candidates [12], lead to skewed evaluation of objective fact and decreased accuracy of opinion [19]. Furthermore, some studies indicate that in some cases these homogeneous sub-networks may reinforce bias [20], in some part due to avoidance of cognitive dissonance [21]. Given that interaction between dislike agents in a network can sometimes correct false beliefs [22], these echo chambers may be seen as drivers of polarisation [23]. This is especially since much anti-vaccine content is shared without thought of its veracity [24], with Facebook anti-vaccine groups serving the dual purpose of opinion-reinforcing echo chamber and “fake news” source in a time where a large number of people draw on social media sites for their health information [25].

In the same vein, much work has focused on the modularity of social networks. Modularity is a graph theoretic measure of the segregation of a graph [26]; a high degree of modularity may indicate increasing segregation of a network into clusters [27], with other work showing that modularity is not a “direct measure of polarisation” [28]. With some governing dynamic, modularity is a “essentially rooted at the stability of its corresponding social system”, with stable networks containing one large community and unstable networks showing modularity driven by polarisation [29]. The global clustering coefficient (GCC) works in a similar way; by describing the number of triangles in the network, it is an important measure of graph structure [30]. A high clustering coefficient is accompanied by the small world phenomenon of short average inter-node distance [31] which facilitates efficient information spread on social networks through redundancy [32]. This clustering also facilitates mutual communication between nodes, where effective communication between disagreeing persons can lead to change of opinion [12].

The occurrence of opinion change within opinion-based communities has seen much attention in political studies, with some work asserting the ability of the ‘wisdom of the crowd’ to overcome bias [33], while other work shows that group phenomena reinforce the opinions held [19]. Others have argued that a commonly held belief within a community can still become more accurate even as the homogeneity of the group increases [34]. This leads to our interest in the rate of opinion change in the network as yet another potential indicator of dynamical regime change. Systems moving from one polarised state to another undergo phase transition through a sole critical point [35]. Called critical transitions, they sometimes result in a demonstration of characteristic system behaviours such as critical slowing down [36]. These events give us easily recognisable ‘hints’ of approaching transitions called early warning signals [37].

We show that trends in all of the measurements described above (modularity, global clustering coefficient, census and sizes of communities and echo chambers) provide early warning signals of epidemic and vaccine crisis events for a coupled disease-behaviour model of childhood disease. We use a binary vaccine opinion dynamic and an *SIRV*_*p*_ disease process occurring on a random network to model a childhood infectious disease. By quantifying and comparing their performance, we find that trends in the sizes of anti-vaccine communities were the best-performing signals, with the modularity and clustering coefficients of communities also performing well. All in all, we verify that changes to fundamental graph structure driven solely by opinion dynamics are good predictors of disease events and aggregate sentiment towards vaccination.

This paper is organised as follows: in Sec. 2, we will describe the disease-behaviour model used in the study and a description of the warning signals used. Section 3 will show the change in trends of the signals with respect to perceived risk of adverse vaccine effects and the social pressure of an injunctive norm, as well as a quantification of the warning provided by each measure through the use of the Standard Normal Homogeneity change point test (SNHT). We will elaborate on the limitations of the measures used and the implication of the results in Sec. 4. Information showing the results obtained through the application of other change point tests are given in the Appendix.

## 2. Methods

### 2.1. The model

For simulation, we use an ABM identical to that described in [38], shown in Fig. 1. A perfectly effective vaccine with no effect on mortality is immediately available to susceptible agents upon gaining pro-vaccine status *S* → *V*_*p*_. As shown in Fig. 1a, the disease follows the *SIRV*_*p*_ model; each agent physically interacts with all their neighbours per time step. If a susceptible agent becomes infected *S* → *I* (with probability 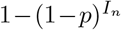, where *I*_*n*_ represents the number of infected neighbours of the agent *n* and *p* = 0.2 the probability of infection), they become both ill and infectious for ℓ = 2 time steps (each time step represents a week); recovery *I* → *R* and vaccination *S* → *V*_*p*_ are both permanent. Injunctive social norms 0 ≤ *σ* ≤ 2 and a cost of 0.4 ≤ *κ* ≤ 0.2 represent peer pressure and adverse vaccine effects respectively [39].

**Figure 1:**
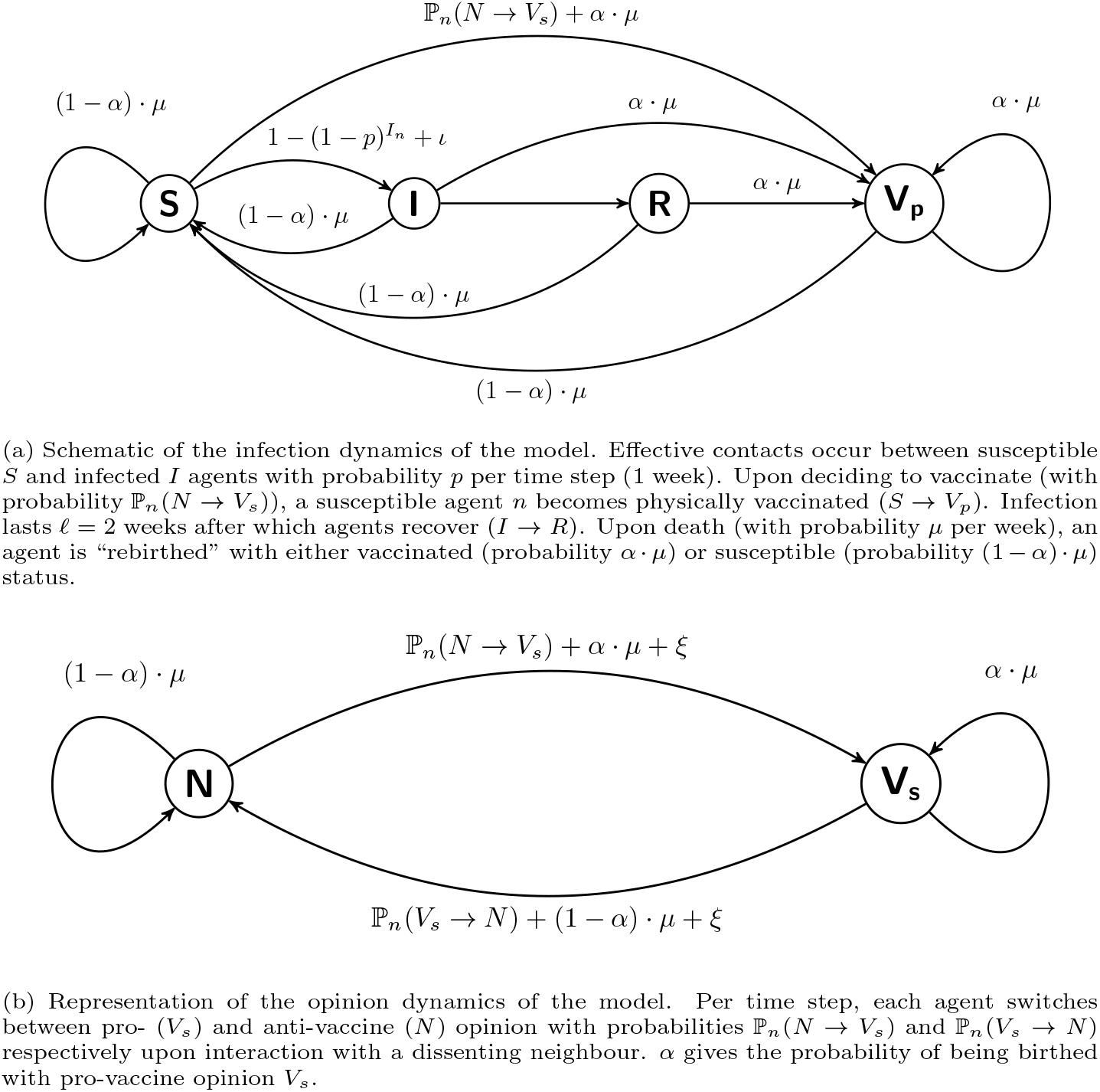
Diagrams showing the physical (a) and social (b) dynamics of model V1.

Figure 1 features a binary social (opinion) dynamic, where agents demonstrate either pro-vaccine (*V*_*s*_) or anti-vaccine (N) sentiment. Change of sentiment occurs through imitation of neighbours, where a randomly chosen neighbour is sampled each time step; an effective interaction with a disagreeing neighbour (with different sentiment than the agent sampling) prompts a reevaluation and change of sentiment with probability ℙ_*n*_(*N*→*V*_*s*_) for anti-vaccine agents and ℙ_*n*_(*V*_*s*_ →*N*) for pro-vaccine agents. Any susceptible agent that adopts pro-vaccine sentiment (*N* → *V*_*s*_) is immediately vaccinated (*S* → *V*_*p*_); for agent *n*, these changes in sentiment depend on the perceived vaccine risk *kappa* and neighbours *I*_*n*_:

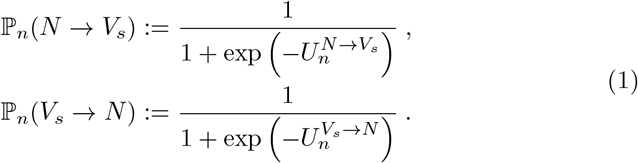

Indices 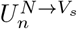 and 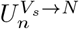 in equation (1) are utility functions defined as

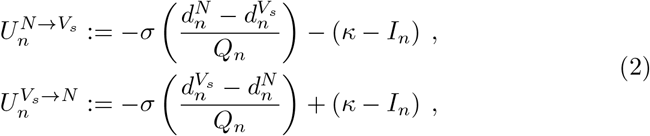

where 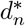 represents the number of neighbours of *n* with sentiment *, with *d*_*n*_ representing the total number of neighbours.

### 2.2. Early warning signals

Four of the six EWS explored in this paper are related to the detection of community structure in networks; the global clustering coefficient, echo chamber, opinion-based community and modularity score are all important tools that enable biological modelling [26]. The topological phenomenon of *community* refers not to a single central construct, but rather a general notion of variation in connection density. Communities in social networks are vaguely defined in the literature as groups with the following basic property: members of the community are more connected to each other than with non-members [40]. Vagueness in science usually leads to artistic licence and the specific treatment of context and purpose; by the above definition, communities can then be alternately conceptualised in different areas and studies as modules, clusters, groups and so on [41]. Such refinements then lead to tighter and more technical definitions.

Here, we conceptualise topological communities as (connected) components on the network. Components in undirected networks are maximal disjoint groups of agents such that there is a path between every pair of agents in the group. [42]. Even more specifically, the concept of a giant connected component (GCC) describes a component that contains a “significant fraction of all the nodes” [43]. The role of components (both non-giant and giant) in spreading processes can be conceptualised as such: where some infection can be spread from person to person, GCCs are formed by historical person-to-person contacts (as opposed to current contacts) [44]. Practical indirect analysis and exploitation of this component structure in policy design and epidemiological intervention is facilitated through contact tracing [43].

Recent political upheaval has thrust the phenomenon of the echo chamber into social consciousness and modern parlance, with many lay articles arguing for and against their existence, though many articles do not directly define the concept before exploiting it. There exist different definitions of echo chambers in literature; also called *tribes* [45], they can be described either as a community where at least some percentage of the members hold a particular sentiment [46], or else a subset of community members overwhelmingly likely to restrict their neighbourhood communication to contacts with shared opinion [47]. As such, echo chambers are usually conceptualised as closed subsystems of social networks containing members with a single orientation [17], and are therefore considered synonymous with homophily and strongly associated with the quick spread of misinformation [48], polarisation and the insulation and reinforcement of belief despite their veracity [17]. One example is the concept of the ‘filter bubble’, detailing the biased filtering of information unfortunately created by personalisation algorithms [49].

Given the described ubiquity on social networks and their importance in information diffusion (and therefore decision making) [48], we test whether observations of the size and number of communities *Z*_*_ and echo chambers *J*_*_ give early warnings of approaching vaccine scares and crises, as well as falls in vaccination rates. We retrieve the echo chambers *J*_*_ by first listing the agents in the network with the desired vaccine opinion; the members of the network are then sorted into two primary classes. The *peripheral* members maintain links with at least one disagreeing neighbour and *core* members only maintain links with agreeing neighbours (these are direct analogies of the boundary and interior of a topological space respectively). We then characterise echo chambers as communities of core members.

Clustering has been shown to greatly facilitate the spread of such phenomena as political extremism [50] and infectious disease [51]. Specifically, clustering refers to the propensity of connection between two persons if they have a mutual friend [52]. As an indicator of this organisation, the *global clustering coefficient* indicates the prevalence of clusters, dense highly-connected groups of nodes in a graph [53]. This done by finding the density of *triplets* on the network. An *open triplet* is a group of three nodes connected by 2 edges, while a *closed triplet* is a group of three nodes joined by three unique edges (also called *triangles* for this reason). The global clustering coefficient (GCC) is then calculated as

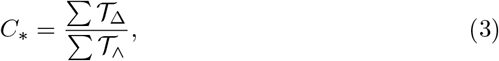

where ∑ 𝒯_Δ_ represents the number of closed triplets and ∑ 𝒯_^_ represents the number of open triplets [54]. The value of the coefficient is bounded 0 ≤ *C*_*_ ≤ 1. In metaphor with percolation theory literature, the clustering coefficient of sub-networks formed by holders of either sentiment could potentially act as an indicator of organisation that enhances the reinforcement of sentiment and the effect of any social norm.

The modularity measure is similar to the global clustering coefficient as a measure of organisation; highly modular networks possess many *modules*, which feature dense interconnectivity between nodes similar in some way and sparse connectivity between dislike nodes. Specifically, this measures the correlation between the probability of connection of two nodes and their membership of the same module [55]. The modularity score is calculated as follows [56]; let an undirected network be divided into two disjoint groups Λ and Ξ, with each node *n* given the score

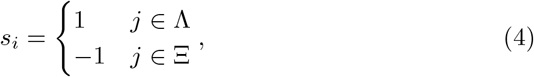

with an adjacency matrix *A* of the network, so that *A*_*ij*_ gives the number of edges between nodes *i* and *j*. Let *k*_*_ represent the degree of node*, so that

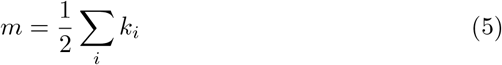

gives the number of edges in the network and the expected number of edges between nodes *i* and *j* is

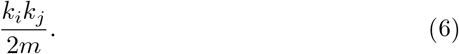

The network modularity (*Q*) is then given as

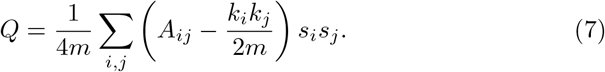

In the model investigated, vaccination occurs only through the first adoption of pro-vaccine opinion *N* → *V*_*s*_ (Sec. 2.1); as such, the number of changes of opinion Θ_*_ undertaken by agents of either opinion can conceivably describe both the social and infection dynamics of the model and thereby yield warning signals of sudden transitions. Moreover, equation (2) shows that the probability of switching sentiment is sensitive to the number of infected agents in the individual neighbourhood; this dependence makes it highly likely that the probability of having an infected neighbor Γ_*_ is correlated not only with vaccine coverage both also with the rate of change of opinion throughout the length of each simulation.

### 2.3. Parameters and time series

Both (infection and social) layers of the network have an Erdős-Rényi random network structure 𝒢 (10000,0.003); network size *N* = 10000 and mean degree ⟨*d*_*n*_ ⟩ = 30 were chosen for alignment with studies of similar coupled behaviour-infection models [57, 38], as well as for computational tractability. Each simulation starts with proportion *α* = 0.25 of vaccinated pro-vaccine agents (states represented by pair (*V*_*s*_, *V*_*p*_)), with all others being susceptible anti-vaccine agents (pairs (*N, S*)). The probability of death per time step is *μ*, so that *α · μ* gives the probability of reset with initial state *V*_*s*_ and (1 − *α*) *μ* the probability of reset with initial state *S*. *ξ* = 1 *×* 10^−4^ represents the probability of switching sentiment randomly. Noise parameter *ξ* = 0.0001 represents unsystematic fluctuations commonly seen in empirical studies [58]. The birth/death probability *μ* = 2.4*×*10^−4^ affords an average life span of 80 years to each agent [59]. The case importation rate *ι* = 2.5 *×* 10^−5^ of susceptible agents adds periodic impulses of infection as a test of resilience in non-endemic disease regimes.

Ensemble size was set at 20 for most parameter combinations used. Figure 2 shows nine time series from the infection and social dynamics for a succession of vaccine risks *κ*; all time series taken over all realisations showed an initial epidemic spread (examples in Figs. 2(a-c)), with subsequent decrease as the pool of susceptible agents is consumed. Noted is that Figs. 2d (*κ* = 0.03125) and 2e (*κ* = 0) both show almost perfect vaccine rates ⟨*V*_*p*_⟩ over 49 realisations of the parameter values, while the corresponding social pro-vaccine consensus shown in Fig. 2g collapses as the perceived vaccine risk*κ*decreases into Fig. 2e. Finally, Figs. 2f and 2i (*κ* = −0.03125) show a median vaccination rate [*V*_*p*_]≈ 0.5 with corresponding anti-vaccine consensus [*V*_*s*_] ≈ 0 respectively. Taken all together, these trends show that simple observation of the social dynamics may not allow for predictions of the infection dynamics and vice versa [38]. This is due to the permanence of the vaccine; an agent can change their vaccine opinion throughout their lifetime, but they cannot become ‘unvaccinated’ should their stances change.

**Figure 2:**
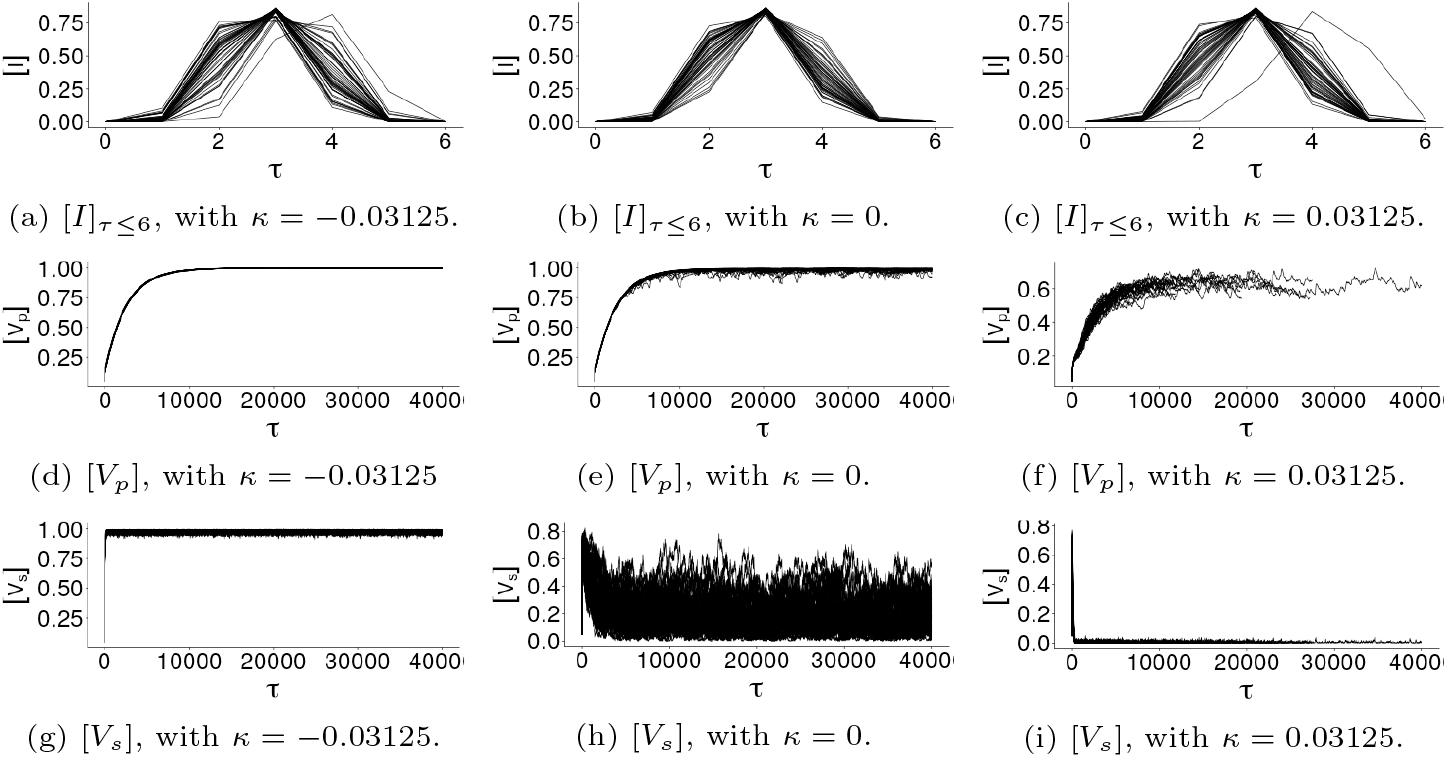
Example time series (49 realisations each) of social (d-f) and infection (g-i) dynamics of the model for social norm *σ* = 0, where time *τ* is measured in weeks (one time step is a week). Row (a-c) shows the initial epidemic spread of the disease over the first 6 time steps of each realisation. The changes in trend of [*V*_*s*_] with increasing vaccination cost *κ* in (g,h) compared to the corresponding graphs of [*V*_*s*_] (d,e) show that the infection dynamics may not be a predictor of the social dynamics and *vice versa*.

The chosen parameter space (*κ,σ*) ∈ [−1,1] *×* [0,3] was sufficiently broad to capture transitions in both the infection (Fig. A.8a) and social (Fig. A.8b) dynamics. The contours in each panel of Fig. (A.8) show the obvious correspondence between social (*K*_*s*_) and infection (*K*_*p*_) transitions and substantial changes in the clustering coefficient of the pro-vaccine sub-network 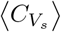 (Fig. A.8c) and the mean size of anti-vaccine communities ⟨|*Z*_*N*_ | ⟩ (Fig. A.8d).

## 3. Results

We again define the *infection transition K*_*p*_ as the value of perceived vaccine risk *κ* value at which the mean number of recovered agents in the model surpasses that of the vaccinated agents and vice versa, so that ⟨*V*_*p*_⟩ ≈ ⟨*R*⟩; in Fig. 3a, *K*_*p*_ is represented by the dashed vertical line marking the approximate intersection of the curves giving ⟨*V*_*p*_⟩ and ⟨*R*⟩. Similarly, the *social transition K*_*s*_ is the *κ* value at which one of ⟨*N*⟩ (number of anti-vaccine agents) and ⟨*V*_*s*_⟩ (number of pro-vaccine agents) surpasses the other so that ⟨*V*_*s*_⟩ ≈ ⟨*N*⟩; this is marked by the dotted vertical line in Fig. 3a showing the approximate intersection of ⟨*V*_*s*_⟩ and ⟨*N*⟩.

**Figure 3:**
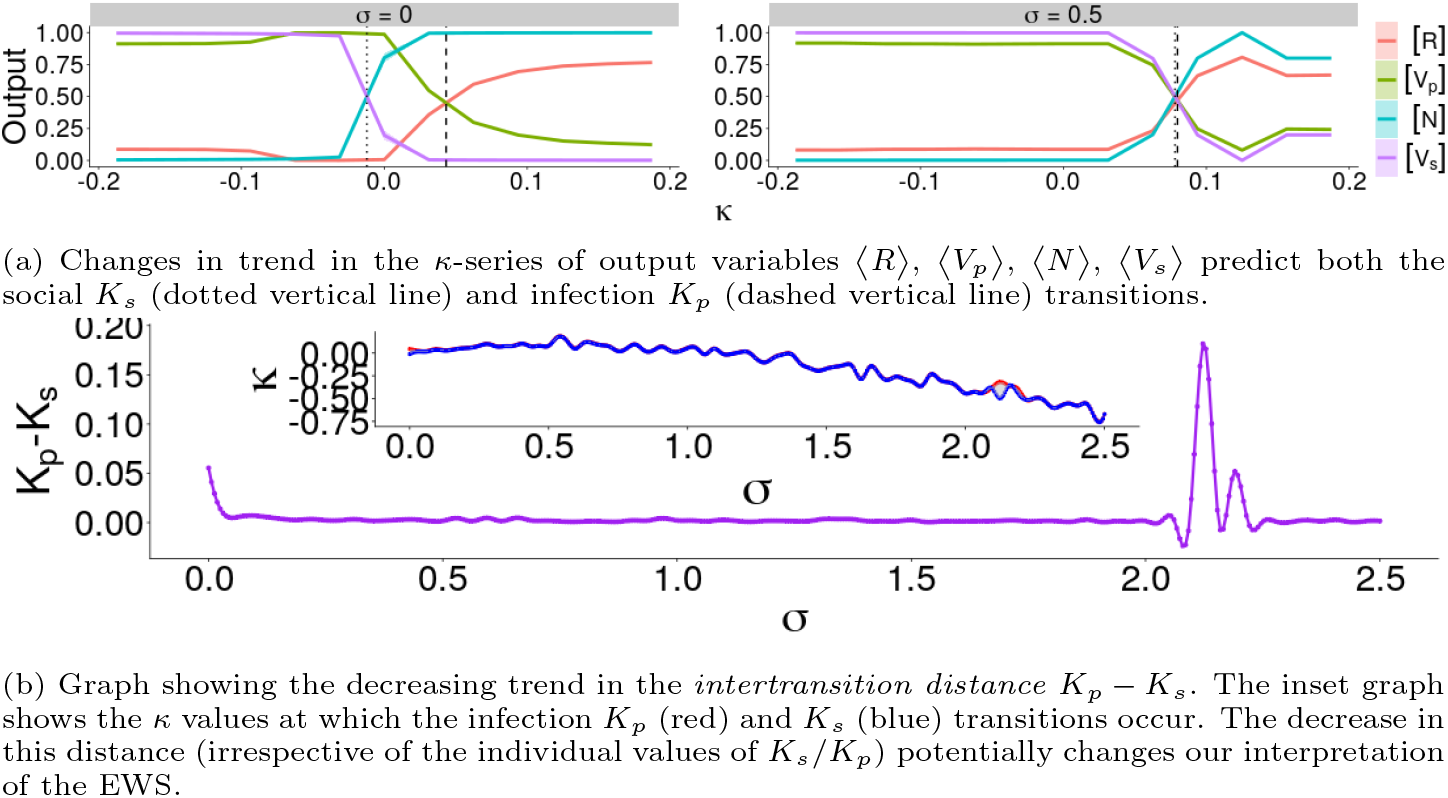
Graphs of the trends in model output variables (with respect to vaccine risk *κ*) and intertransition distance *K*_*p*_ −*K*_*s*_ (with respect to the social norm *σ*).

The distance between the two transitions (i.e.*K*_*p*_ − *K*_*s*_) is called the *inter-transition distance*[38]; as with similar models, we find that this intertransition distance decreases with increasing strength of the social norm *σ*. This is explicitly demonstrated by Fig 3a, where an increase in the social norm from *σ* = 0 (Fig. 3a, left) to *σ* = 0.5 (Fig. 3a, right) brings the two vertical lines (representing transitions *K*_*s*_ and *K*_*p*_ respectively) together.

Figure 3b gives a full picture of the intertransition distance over the investigated parameter range 0 ≤ *σ* ≤ 2.2; its decreasing trend with strengthening social norm *σ* is shown by the purple curve, with the inset panel showing the locations of the social *K*_*s*_ (blue) and infection *K*_*p*_ (red) transitions. The positivity of the graph tells us that *K*_*s*_ *< K*_*p*_ for all strengths of the social norm *σ* ≤ 2.5, so that the social transition always precedes the disease transition. Since physical vaccination is driven primarily by changes in sentiment, this is to be expected; strengthening social norms increases the alignment of behaviour and vaccine uptake, resulting in decreasing intertransition distance *K*_*p*_ − *K*_*s*_.

Warning signals can potentially indicate both transitions, or maybe only one of the two; this subtlety is lost with shrinking intertransition distance and so may impact the predictive power of any early warning signals tested.

### 3.1. Group size and census predict the social transition

Figure 4 shows the trends in the means of the numbers and sizes of communities and echo chambers (both pro- and anti-vaccine) at equilibrium. As for the number of echo chambers ⟨#*J*_*_⟩ (Fig. 4d), there is not much resemblance to the trend of the mean number of connected components ⟨#*Z*_*_⟩ (Fig. 4b); this is partly due to the low occurrence of echo chambers in this network. For *σ* = 0 (Fig 4d, left), there is no warning given by the mean number of pro-vaccine echo chambers ⟨#*J*_*V*_⟩ ≈ 0, whereas the mean number of anti-vaccine echo chambers ⟨#*J*_*N*_⟩ increases before *K*_*s*_, reaching a maximum between the two transitions and reaching zero value approaching *K*_*p*_. At first glance, the change in trend as the social norm strengthens to *σ* = 0.5 (Fig 4d, right) seems substantial.

**Figure 4:**
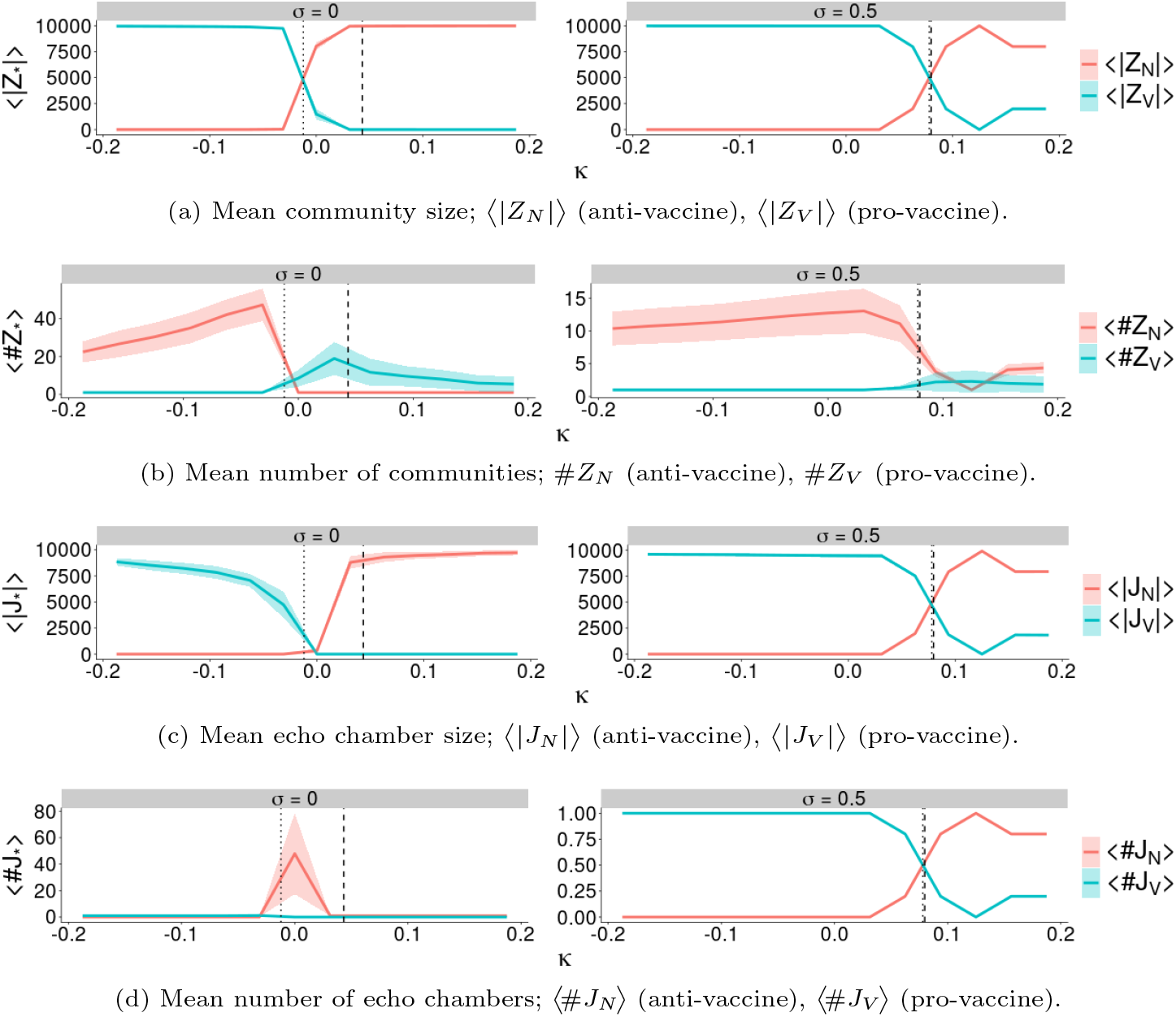
Trends of some measures of connectivity of the social network with respect to the perceived risk of vaccination *κ*. Vertical dashed and dotted lines representing the infection (*K*_*p*_) and social (*K*_*s*_) transitions (respectively) help to illustrate changes in the trends as transitions are approached. The strength of the social norm is *σ* = 0 for panels on the left and *σ* = 0.5 for panels on the right.

Another view of the social dynamics involves analysing pairs of graphs. For example, Figs. 4a and 4b (*σ* = 0) together show the existence of a large community of pro-vaccine agents; for *κ* = −0.25, there is a single community of pro-vaccine agents with size ≈ 9966.56,

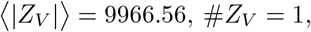

and a few triads of anti-vaccine agents (⟨|*Z*_*N*_|⟩ ≈ 3.09,#*Z*_*N*_ ≈ 19.5). This indicates the existence of one large pro-vaccine component interspersed with small triads of anti-vaccine agents. **H**owever, when vaccine risk increases to *κ*= 0.25, there is a single anti-vaccine community of mean size 9992.8

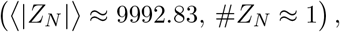

with a few individual pro-vaccine agents (⟨|*Z*_*V*_|⟩ = 1.82, ⟨#*Z*_*V*_⟩ = 4.34). These observations hold for the stronger social norm *σ* = 0.5 in Figs. 4a and 4b.

Intuitively, a similar pattern holds for the formation and erosion of echo chambers seen in Figs. 4c and 4d (*σ* = 0), and 4c and 4d (*σ* = 0.5). The scenario ⟨*V*_*s*_⟩ ∼ ⟨*N*⟩ (for small *κ*) necessarily leads to the creation of a large community of pro-vaccine agents and thereby a large echo chamber; conversely, ⟨*V*_*s*_⟩ ≈ 1 will result in small dispersed components of pro-vaccine agents with a small or empty interior, resulting in a very low number of echo chambers.

We use the Standard Normal Homogeneity Test (SNHT) from the `trend` package in R [60] to find a change point (SNHT{*}) in the *κ*-series of each EWS for each strength of the social norm *σ*; the distance between the transition *K*_*s*_ and the change point is called the *lead distance*. Each panel of Fig. 5 gives the trends of lead distance *K*_*s*_ − SNHT{*} of each EWS on the left, and a bar chart on the right showing the number of *σ* values for which each EWS gave the highest lead distance measured; positive trends indicate that the warning precedes the social transition. Each panel of Fig. 5 shows that most of the EWS shown give positive lead distances for *σ* ≤ 2.875, and that most lead distances decrease as social norm *σ* strengthens.

**Figure 5:**
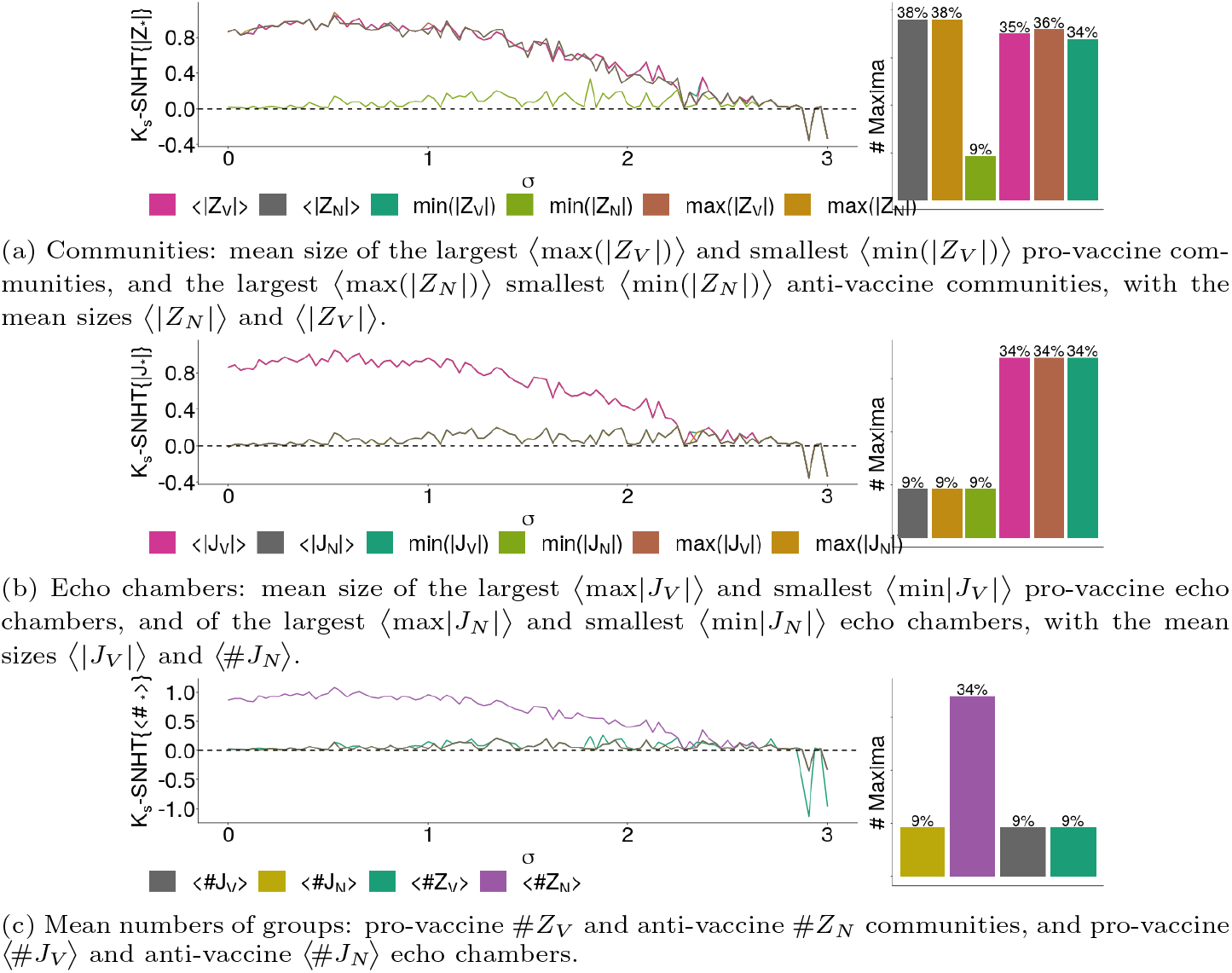
Trends in the lead distance *K*_*s*_ − SNHT{*} for each EWS’*κ*-series with respect to the strength of the social norm *σ*. The bar chart on the right of each panel shows the percentage of *σ* values for which the measured EWS gave the largest lead distance of all the EWS tested.

Figures 5b and 5c expose all measurements of anti-vaccine echo chambers *J*_*N*_ as bad performers, along with the size of the smallest anti-vaccine community ⟨min(|*Z*_*N*_ |) ⟩ (Fig. 5a), the number of pro-vaccine communities ⟨#*Z*_*V*_ ⟩ (Fig. 5c) and the number of pro-vaccine echo chambers ⟨#*J*_*V*_ ⟩ (Fig. 5c); they all give very little warning of the social transition *K*_*s*_ for all strengths of the social norm *σ*, as well as giving the best warning (of all the EWS) for only ≈9% of the tested range of the social norm *σ*. They all demonstrate significantly lower lead distances than those of the other EWS, giving the highest lead distance of all EWS for only 9% of *σ* values (as compared to ≥30% for other EWS shown in Fig. 5). The performance of these EWS under different change point detection tests is shown in Figs. (C.11-C.13). As social norm *σ* increases, pressure on each agent to conform to surrounding opinion becomes the main driver of self-organisation of the social dynamics. Since it also increases the speed of this transition between these opposing organised states, lead distances *K*_*s*_ −SNHT{*}will decrease as the social morn *σ* increases.

### 3.2. Clustering and uncertainty also provide early warnings

Figure (B.9) shows the trends in other EWS with respect to the social norm *σ*; modularity of the social network ⟨*Q*_*_⟩, the global clustering coefficient ⟨*C*_*_⟩, number of sentiment changes ⟨Θ_*_⟩ and the probability of having an infected neighbour ⟨Γ_*_⟩. Graph modularity ⟨*Q*_*_⟩ and the number of sentiment changes ⟨Θ_*_⟩ give appreciable warning signals both transitions in Fig. (B.9), given their noticeable changes in trend approaching *K*_*s*_ (dotted vertical line) and *K*_*p*_ (dashed vertical line). However, the likelihood of having an infected neighbour ⟨Γ_*_⟩ only predicts the disease transition when *σ* = 0; the probability of having an infected neighbour for both pro-vaccine ⟨Γ_*V*_⟩ and anti-vaccine ⟨Γ_*N*_⟩ in Fig. (B.9c) (for social norm *σ* = 0) only show substantial visible changes in trend approaching the disease transition *K*_*p*_, unlike its uneventful approach to the social transition *K*_*s*_.

This behaviour disappears when social norm *σ* →0.5 (Fig. B.9c, right) due to the shrinking intertransition distance *K*_*p*_ −*K*_*s*_; lead distances *K*_*p*_ – SNHT{⟨Γ_*_⟩} of both probabilities ⟨Γ_*N*_⟩ and ⟨Γ_*V*_⟩ for *σ* = 0 and *σ* = 0.5 are similar (0.95 and 0.98 respectively), showing that the distances between the warnings and the transitions *K*_*p*_ are similar in both cases. As *σ* increases to 0.5, *K*_*s*_ ‘moves closer’ to *K*_*p*_; this suggests that any warning of *K*_*s*_ occurs incidentally, rather than being directly caused by the model dynamics. This is intuitive; any measure of the probability of having an infected neighbour is an observation of the infection dynamics, therefore an assumption of some direct relationship between ⟨Γ_*_⟩ and *K*_*p*_ is natural.

In Figs. (B.9d), the trends of the global clustering coefficient ⟨*C*_*_⟩ are dwarfed by their standard deviations; Figs. (B.9e) shows the trend without indicating the standard deviation of the mean, for clarity. In Figs. (B.9d) and (B.9e), the respective clustering coefficients of the sub-networks formed by only anti-vaccine (⟨*C*_*N*_ ⟩) and pro-vaccine (⟨*C*_*V*_⟩) agents both show changes in trend approaching both *K*_*s*_ and *K*_*p*_. The global clustering coefficient of the entire social network ⟨*C*_Σ_⟩ holds constant value ≈0.03, leading it to have the second-worst performance of all the EWS tested (Fig. 6).

**Figure 6:**
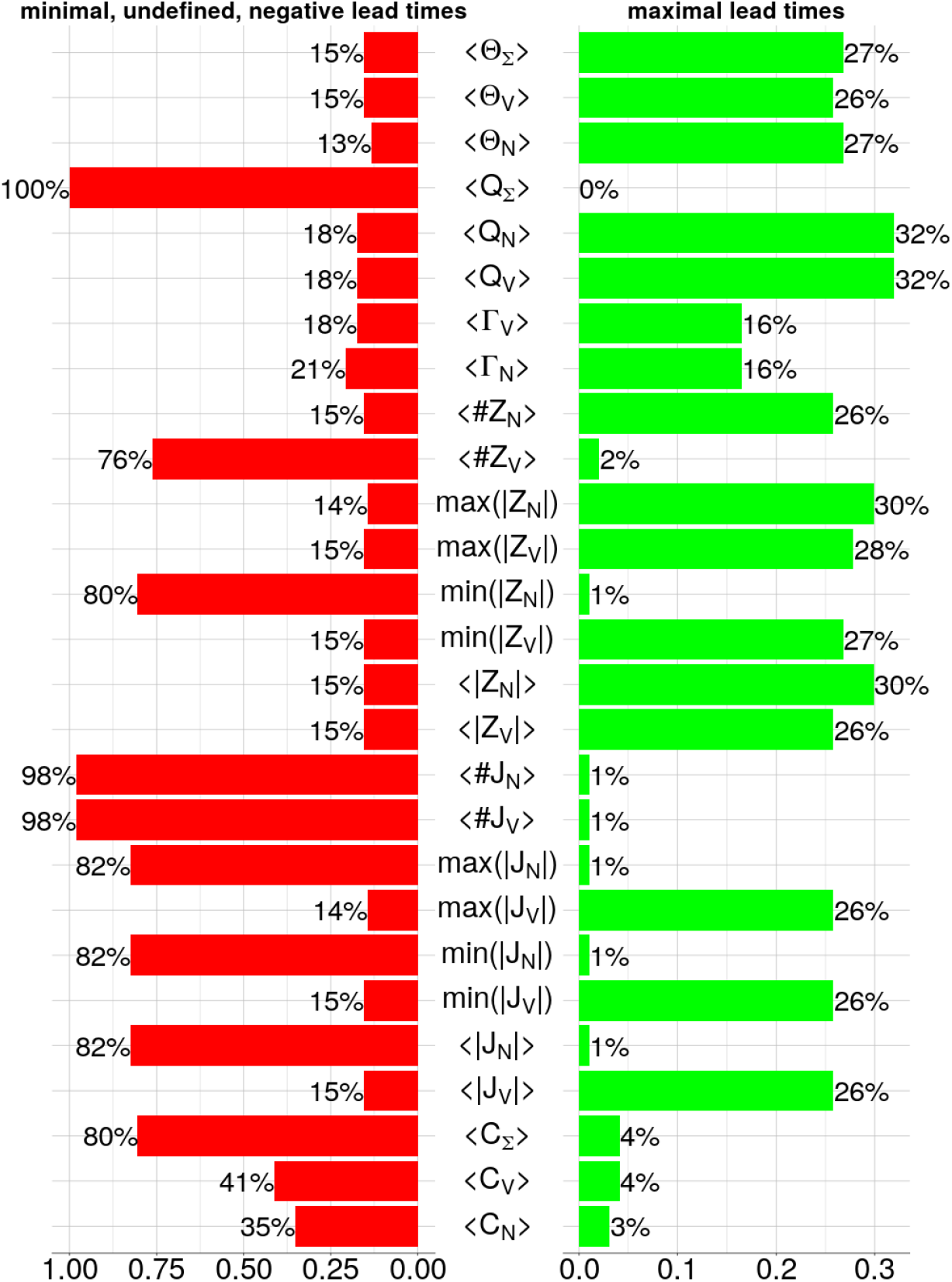
The proportions of social norm *σ* values for which each EWS gives the greatest (green) and least (red) warnings (biggest and smallest lead distances), corresponding to the charts shown in each panel of Figs. 5 and (B.10).

Figure (B.10) shows the trends in lead distances of the remaining EWS. For all the EWS in Fig. (B.10), we can see that lead distance decreases with strengthening social norm *σ*, with all the EWS eventually failing (giving negative lead distances, so that warnings *follow K*_*s*_ - these are useless); modularity scores for the sub-networks formed by pro-vaccine ⟨*Q*_*V*_⟩ (Fig. B.10a) and anti-vaccine ⟨*Q*_*N*_⟩ (Fig. B.10a) agents give useful warnings for all social norms *σ* ≤ 2.40625, while both the number of opinion changes ⟨Θ_*_⟩ (Fig. B.10b) and the probability of having an infected neighbour SNHT{⟨Γ_*_⟩} give useful signals for *σ* ≤ 2.90625. As stated, the global clustering coefficient of the entire social network SNHT{⟨*C*_Σ_⟩}(Fig. B.10d) was undefined for most *σ* and negative for quite a few others, resulting in the worst performance of all the EWS tested and giving the highest lead distance for only 4% of the total range of the social norm *σ*. The sub-networks generated by pro-vaccine agents gave often unsubstantial though positive leads SNHT{⟨*C*_*V*_⟩} (Fig. B.10d), while the sub-networks formed by anti-vaccine agents (Fig. B.10a) gave very little lead distance over most of the range of *σ*. The performances of these EWS under different change point detection tests are shown in Appendix C.

### 3.3. Finding the best and the worst EWS

We compare the EWS by finding the proportion of *σ* values for which each EWS gives the largest warning; this is shown in Fig. 6. We specify a *good* warning as one that gives the *highest* lead distance of all warnings for a single *σ* value and a *bad* warning as one that gives either a *minimal, negative or undefined* lead distance. For many *σ* values, the largest and smallest lead distances were not unique, so that the ratios on neither side sum to 100%.

Comparison of the panels of Fig. 6 gives a notion of ‘dependability’; were a restricted set of EWS to be employed, we would prefer *good* EWS (ones that give the largest lead distances) that aren’t also *bad*(giving the smallest lead distances). Some of the foremost EWS best satisfying this criterion are the counts of all different opinion changes ⟨Θ_*_⟩ and the mean size of anti-vaccine communities ⟨|*Z*_*N*_|⟩ (30% best, 15% worst). The best performers are the modularity scores of the opinion sub-networks ⟨*Q*_*V*_⟩, being the best EWS for 32% of *σ*values, with 18% bad warnings. Conversely, Fig. 6 suggests that the numbers of echo chambers ⟨#*J*_*_⟩ both bring up the rear, providing good warnings for only 1% of *σ* values while giving bad warnings for 98% of social norm *σ* values tested.

Overall, all observations of the sizes of *anti*-vaccine echo chambers |*J*_*N*_| on the network are poor EWS. Conversely, the sizes of *pro*-vaccine echo chambers |*J*_*V*_ |perform generally well (≥26% best, 15% worst). Also, the mean and maximum sizes of anti-vaccine communities (⟨|*Z*_*N*_|⟩ and ⟨max(|*Z*_*N*_|)⟩ respectively) give two of the highest ratios of best warnings (both 30%) and not many bad warnings (≤15%), but observations of the minimum size ⟨min(|*Z*_*N*_|)⟩ give only 1% good warnings and 52% bad warnings. The global clustering coefficient ⟨*C*_*_⟩ performed particularly badly as an EWS, with ≤ 4% best and 35% − 80% worst warnings.

Another observation made from Fig. 6 would be the relationship between the percentages of good and bad warnings; they are actually strongly anti-correlated, with a coefficient of −0.77. The previously shown behaviours of the lead distances eliminate the possibility of an EWS that densely alternates between good and bad warnings, but some EWS do neither; for example, the total modularity of the network ⟨*Q*_Σ_⟩ is trivially neither good (0%) nor bad (0%) by virtue of being everywhere undefined. Nontrivially, the clustering coefficient of the anti-vaccine sub-network ⟨*C*_*N*_⟩ gives intermediate warnings (neither good nor bad) for 60% of social norm strengths *σ*. Therefore, no choice need be made between minimising the percentage of bad warnings and maximising the percentage of good warnings; both strategies yield largely identical results.

Figure 7 show the performance of each EWS per *σ* value. Red tiles show where the EWS gave the smallest positive lead distance of all its peers, while green tiles represent the *σ* values for which the EWS gave the largest lead distance. Yellow columns show where all EWS gave equal lead distances and black tiles represent failed warnings (either undefined or negative lead distances). Therefore, the relative length of an EWS’ red bar in Fig. 6 represents the percentage of that EWS’ red tiles in its row in Fig. 7; the same correspondence holds between the length of an EWS’ green bar in Fig. 6 and the percentage of green tiles in the related row in Fig. 7.

**Figure 7:**
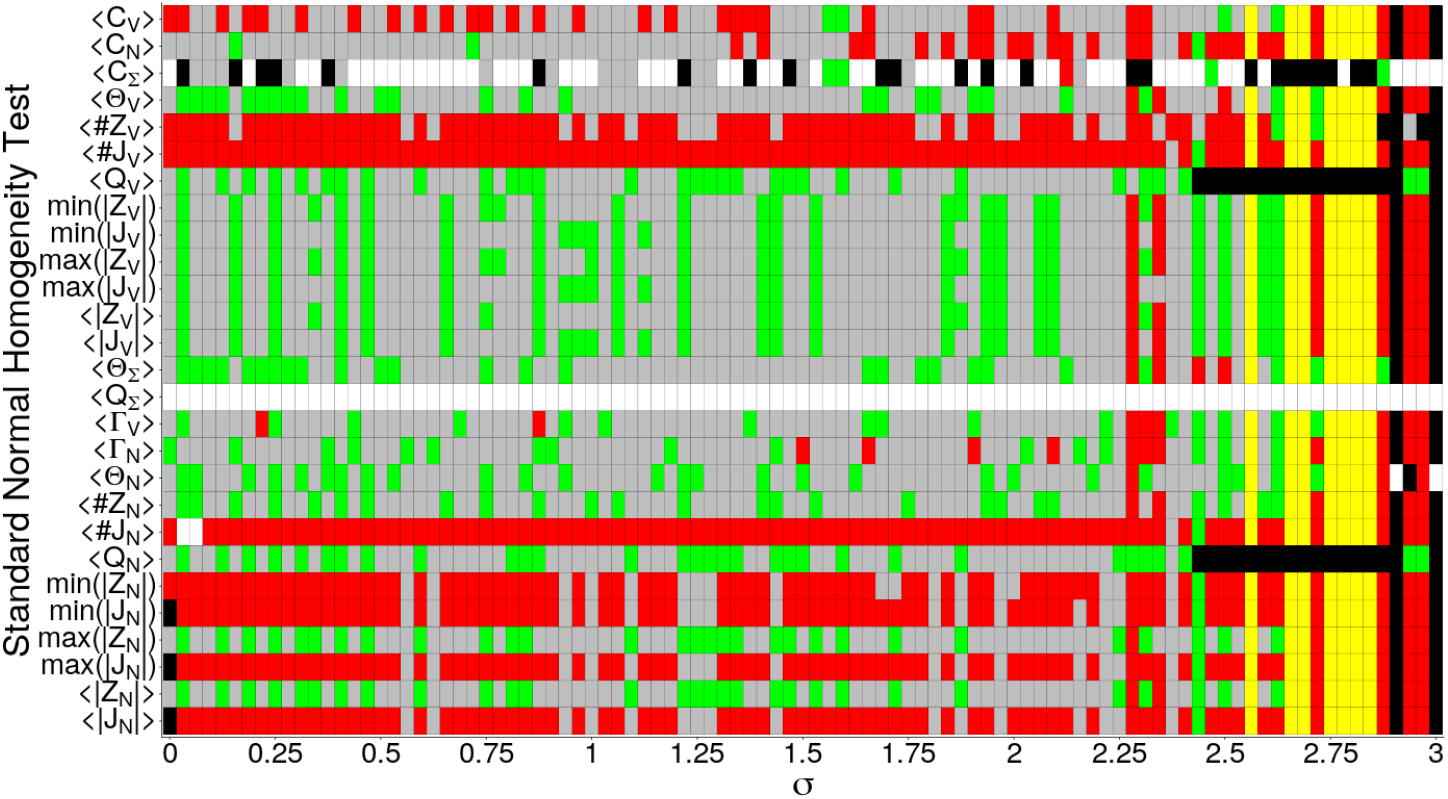
Grid showing the relative performance of each EWS. Green tiles denote the *σ* values for which the EWS gave the highest lead distance, red tiles represent the smallest lead distance, grey tiles represent lead distances that are neither maxima nor minima and yellow tiles show where all EWS gave the same lead distances. Black squares represent failed warnings (negative lead distances) and white tiles represent undefined values (no lead distance).

The main insight provided by Fig. 7 deals with patterns of performance; for instance, the overwhelming red colouration of the rows corresponding to the mean number of pro-vaccine communities and anti-vaccine echo chambers (⟨#*Z*_*V*_⟩, ⟨#*J*_*N*_⟩ respectively) and all EWS related to anti-vaccine echo chambers *J*_*N*_ show that these EWS are quantitatively the worst of the group. Also, there is no detectable pattern in performance visible on the grid; in other words, the effectiveness of the EWS cannot be broken down by ranges of *σ* value. For higher values of the social norm *σ* ≥ 2.625, the prevalence of yellow columns provides the observation that performance seems not to vary as much among the EWS as it does for smaller *σ* values, but nothing else is immediately apparent.

For social norms *σ* = 2.90625 and *σ* = 3, none of the EWS give valid warnings; lead distances are all negative, except for the total clustering coefficient ⟨*C*_Σ_⟩, the number of opinion changes by anti-vaccine agents ⟨Θ_*N*_⟩ and the network’s total modularity score ⟨*Q*_Σ_⟩ which are undefined. This confirms behaviour seen for large *σ* in Figs. 5 and (B.10); indeed, ⟨*C*_Σ_⟩ is undefined for most social norms *σ* because of the disconnection in the social network. Total modularity ⟨*Q*_Σ_⟩ is everywhere undefined (row of white tiles) for this reason.

## 4. Conclusion

In this paper, we tested the use and effectiveness of different network measures as early warning signals (EWS) of sudden transitions in the social and infection dynamics of a multiplex model of disease. For the parameter values used, we found that observations of the mean and maximum sizes of anti-vaccine communities appear to be the most effective EWS of all tested, unlike the size of the smallest anti-vaccine community (though it does give warnings signals); trends in the global clustering coefficient of the sub networks formed by pro- and anti-vaccine agents respectively also marked these events, as well as the number of both respective communities and echo chambers preceding both transitions. This reflects the breakup of connected components on a network preceding a critical transition, an observation well supported by literature on percolation thresholds in random graphs.

A phenomenon of particular interest in this study was the formation and breakup of pro- and anti-vaccine echo chambers; we found that all observations of the sizes of *pro*-vaccine echo chambers (maximum, minimum and mean) performed well as warning signals, while observations of the sizes of *anti*-vaccine echo chambers performed poorly compared to other EWS. The modularity measure of the social network and the rate of opinion changes also warn of transitions of the social and disease transitions, representing changes in aggregate vaccine opinion and vaccine uptake crises respectively. As a direct observation of the infection dynamics of the model, the probability of having an infected neighbour (for both pro- and anti-vaccine agents) performed well as an EWS of vaccine crisis.

Through our proposal and study of effective graph connectivity measures, this study complements others in the field of early warning signals. A potential limitation to the study is our strict definition of an echo chamber; it remains to be seen whether different descriptions such as those featured in other studies of social media networks will result in a more effective or dependable EWS. Also, the graph connectivity measures seem suitable for tracking the dynamics of an evolving network; the inclusion of preferential link formation in the dynamics, as well as social and ‘on the ground’ interventions for different strengths of the social norm, present other interesting avenues of research. Finally, the inclusion of directionality of communication in the network may render the model more realistic [61, 62, 63, 64].

Together with other markers of spatial correlation and aggregation, the graph connectivity measures presented here contribute to the set of tools allowing us to leverage the ubiquity of social media involvement and the resulting data sets in the pursuit of adaptive strategies for maintaining public health.

## Data Availability

Available upon reasonable request.

## Notes

### Competing Interest Statement

The authors have declared no competing interest.

### Funding Statement

This research was funded by an NSERC Discovery Grant to CTB.

