## Appendices for "Echo chambers as early warning signals of widespread vaccine refusal in social-epidemiological networks"

### Supplementary Information.

#### Appendix A. Contour plots of the parameter space

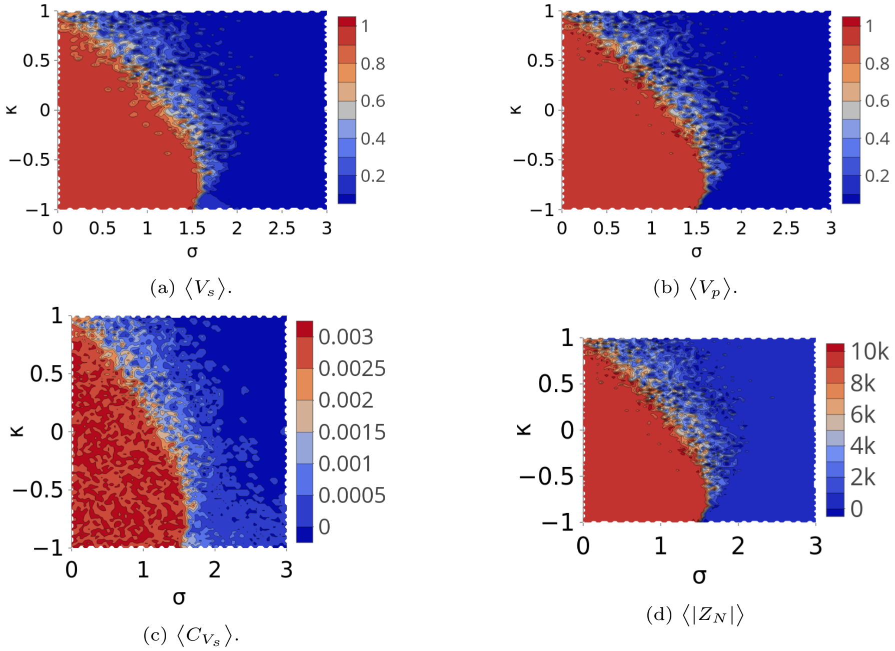


(a) $\langle V_s \rangle$.

(b) $\langle V_p \rangle$.

(c) $\langle C_{V_s} \rangle$.

(d) $\langle |Z_N| \rangle$

Figure A.8: Contour plots of the region $(\sigma, \kappa) \in [0, 3] \times [-1, 1]$ of the parameter plane, capturing the transition dynamics of both the social and infection dynamics; $\sigma$ represents the strength of injunctive social norms and $\kappa$ the perceived health risk of the vaccine. The probability of infection is $p = 0.2$, with $\langle |Z_N| \rangle$ the mean size of anti-vaccine communities and $\langle C_{V_s} \rangle$ the clustering coefficient of the sub-network of pro-vaccine agents.

### Appendix B. Clustering and uncertainty also provide early warnings

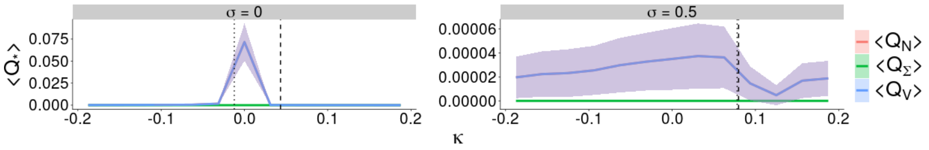


(a) Network modularity. $\langle Q_N \rangle$ (anti-vaccine), $\langle Q_V \rangle$ (pro-vaccine), $\langle Q_\Sigma \rangle$ (total).

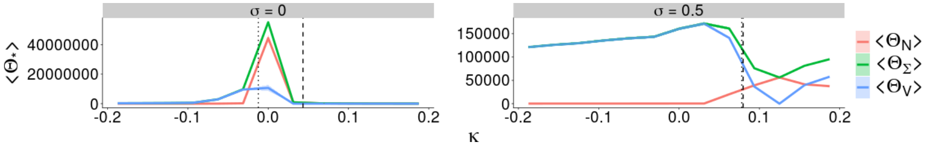


(b) Changes of opinion. $\langle \Theta_N \rangle$ (anti-vaccine), $\langle \Theta_V \rangle$ (pro-vaccine), $\langle \Theta_\Sigma \rangle$ (total).

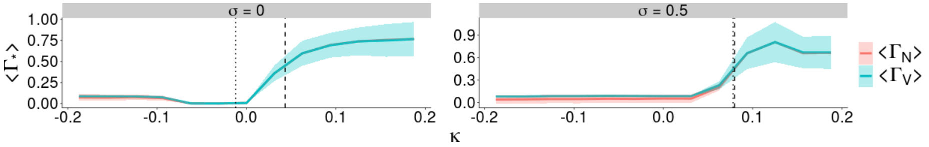


(c) Probability of infected neighbours. $\langle \Gamma_N \rangle$ (anti-vaccine), $\langle \Gamma_V \rangle$ (pro-vaccine).

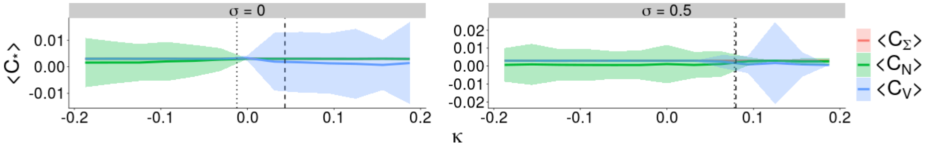


(d) Clustering coefficient. $\langle C_N \rangle$ (anti-vaccine), $\langle C_V \rangle$ (pro-vaccine), $\langle C_\Sigma \rangle$ (total).

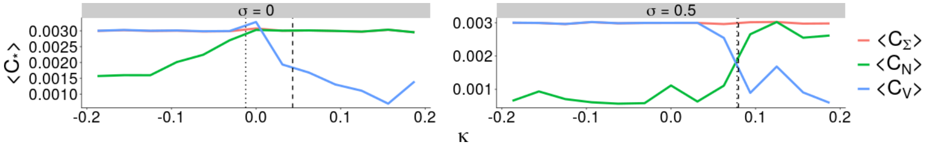


(e) Clustering coefficient (no ribbon). $\langle C_N \rangle$ (anti-vaccine), $\langle C_V \rangle$ (pro-vaccine), $\langle C_\Sigma \rangle$ (total).

Figure B.9: Trends of connectivity measures with respect to perceived vaccine risk $\kappa$. Vertical dashed and dotted lines represent the disease ($K_p$) and social ($K_s$) transitions (respectively). (e) shows the trends of the clustering coefficient without any indications of the standard deviation of the mean, for clarity. Social norm $\sigma = 0$ for the panels on the left, and $\sigma = 0.5$ for panels on the right.

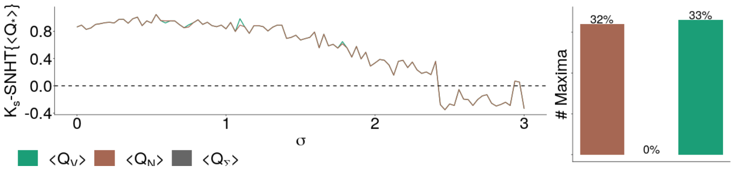


(a) Network modularity score of pro- and anti-vaccine sub-networks.

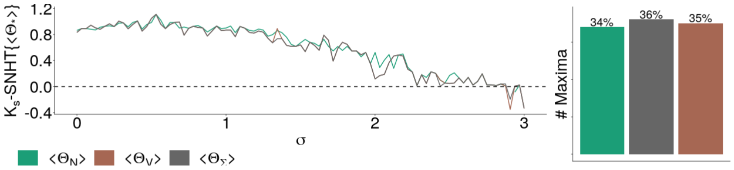


(b) Number of sentiment changes experienced by pro- and anti-vaccine agents.

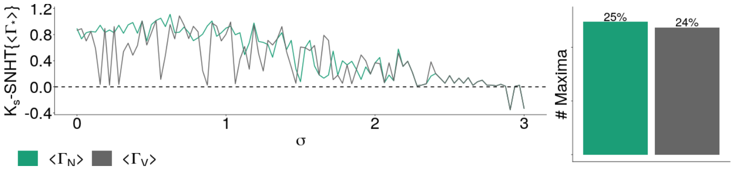


(c) Probability of a pro- and anti-vaccine agents having an infected neighbour.

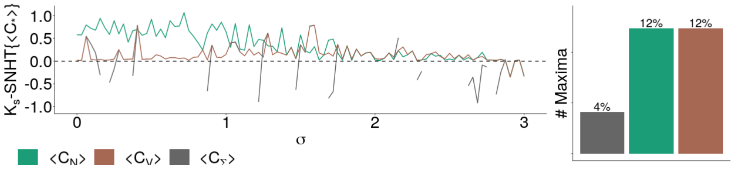


(d) Global clustering coefficient of pro- and anti-vaccine sub-networks.

Figure B.10: Trends in the lead distances of the remaining EWS with respect to the strength of the social norm $\sigma$. Compared to the other metrics shown here, the global clustering coefficient SNHT$\{\langle C_* \rangle\}$ performs badly.

#### Appendix C. Lead time plots

The figures in this section show the lead times given by various EWS with respect to the social norm $\sigma$ with different disease infectivities $p$ and the four different change point tests. In all figures: panels (a) represent the standard normal homogeneity test (SNHT), panels (b) represent the Lanzante test, panels (c) the Pettitt test and panels (d) the Buishand range test. Left panels give results for disease infectivity $p = 0.2$, while panels on the right represent infectivity $p = 0.8$.

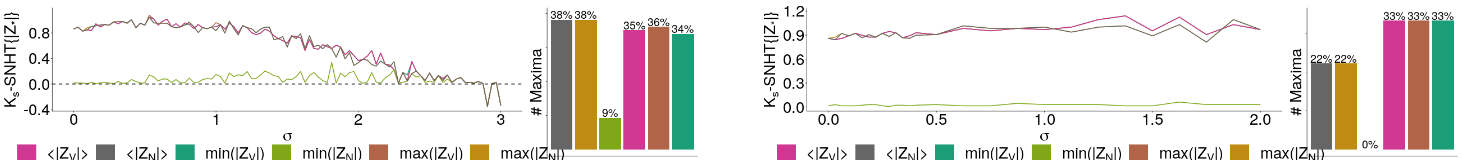


(a) Standard normal homogeneity test.

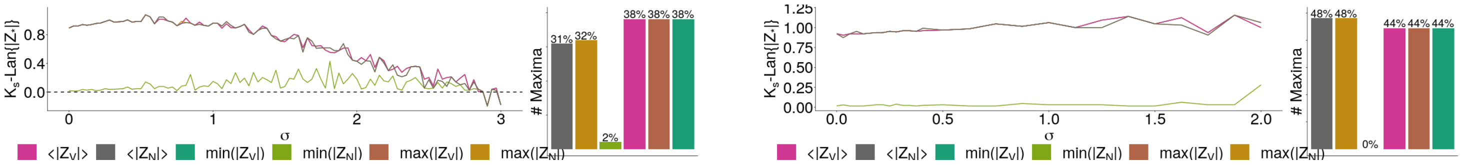


(b) Lanzante test.

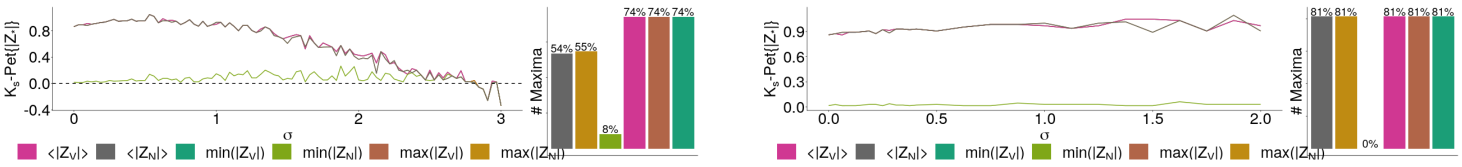


(c) Pettitt test.

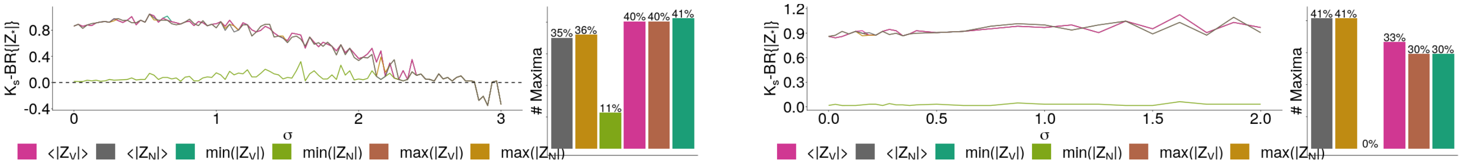


(d) Buishand range test.

Figure C.11: Trends in the lead times of the sizes of different types of opinion communities ($|Z_*|$) under different change point tests. Left panels give results for disease infectivity $p = 0.2$, while panels on the right represent infectivity $p = 0.8$.

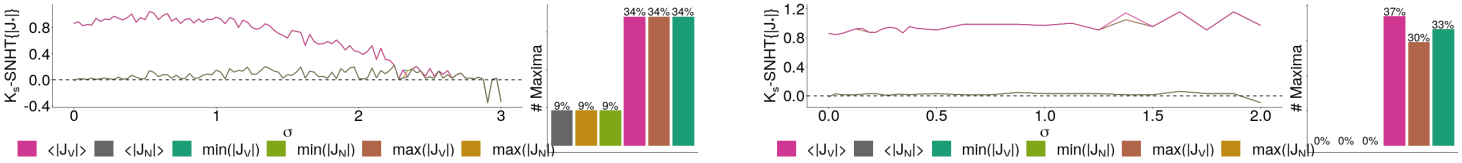

(a) Standard normal homogeneity test.

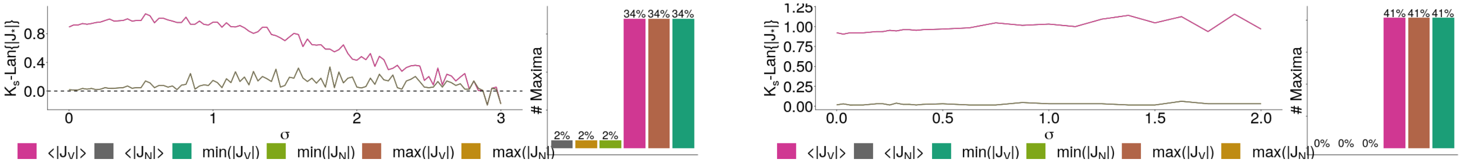

(b) Lanzante test.

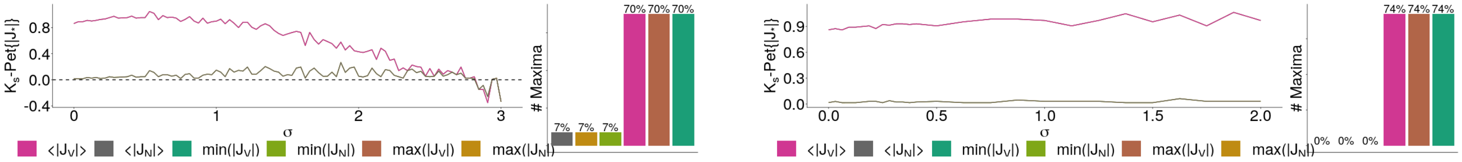

(c) Pettitt test.

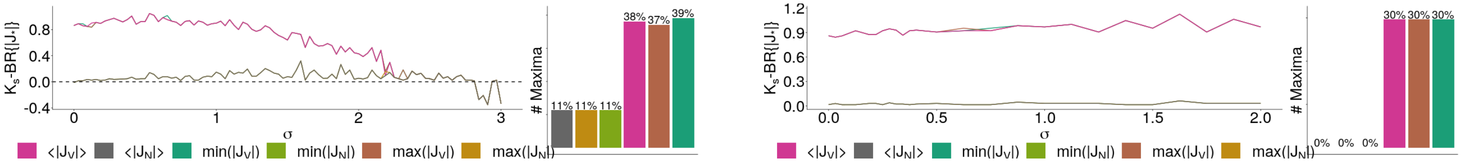

(d) Buishand range test.

Figure C.12: Trends in the lead times of the sizes of different types of echo chambers ($|J_*|$) under different change point tests. Left panels give results for disease infectivity $p = 0.2$, while panels on the right represent infectivity $p = 0.8$.

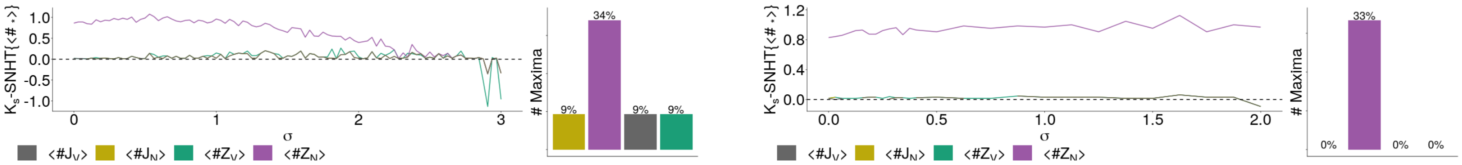


(a) Standard normal homogeneity test.

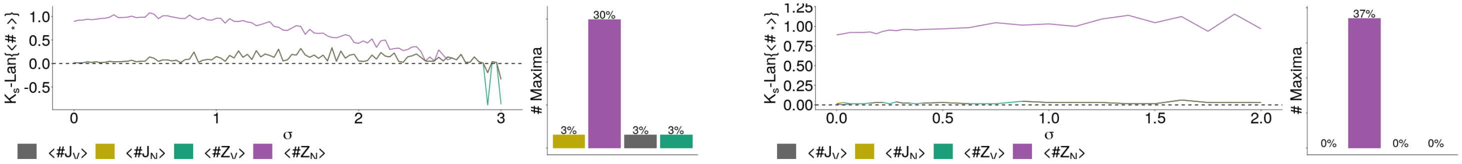


(b) Lanzante test.

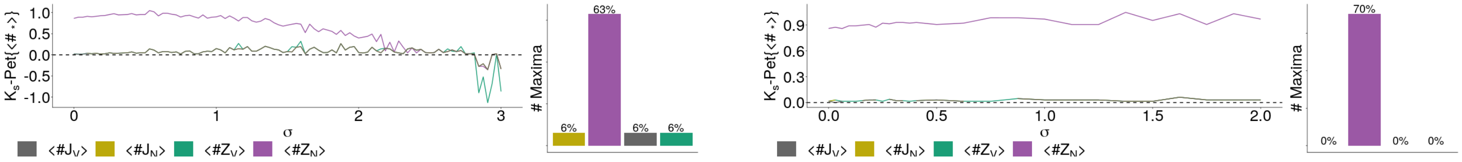


(c) Pettitt test.

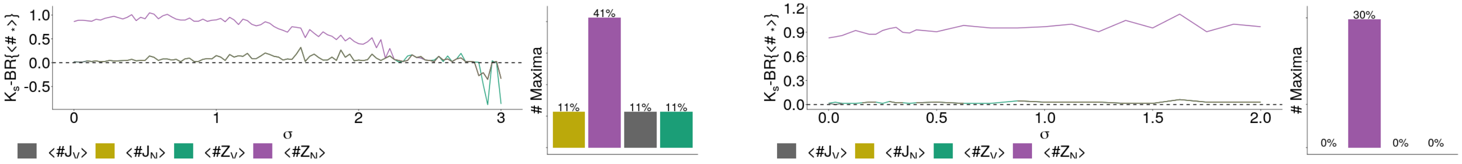


(d) Buishand range test.

Figure C.13: Trends in the lead times of the censuses of different opinion communities ($\#Z_*$) under different change point tests. Left panels give results for disease infectivity $p = 0.2$, while panels on the right represent infectivity $p = 0.8$.

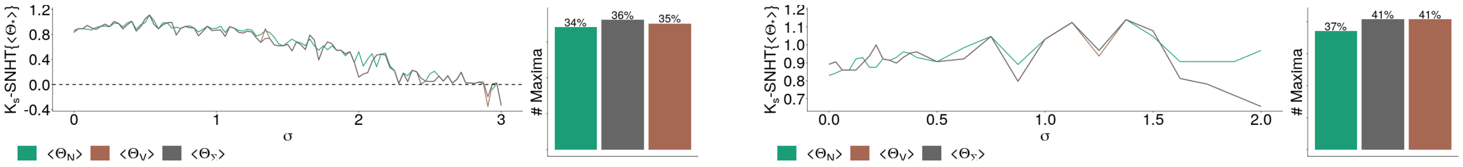


(a) Standard normal homogeneity test.

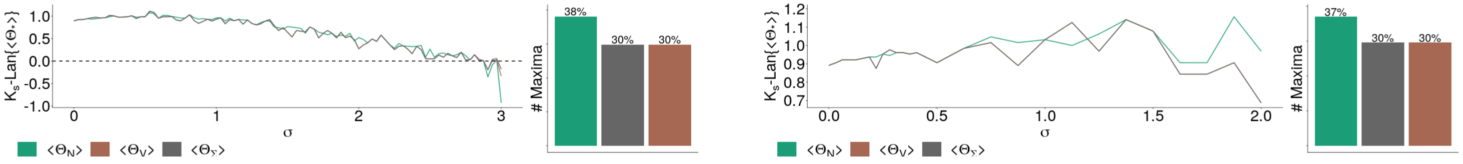


(b) Lanzante test.

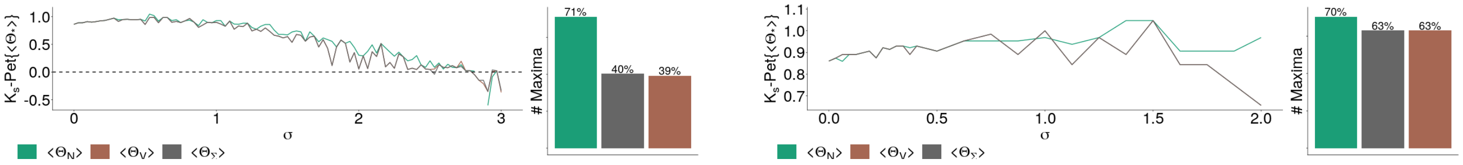


(c) Pettitt test.

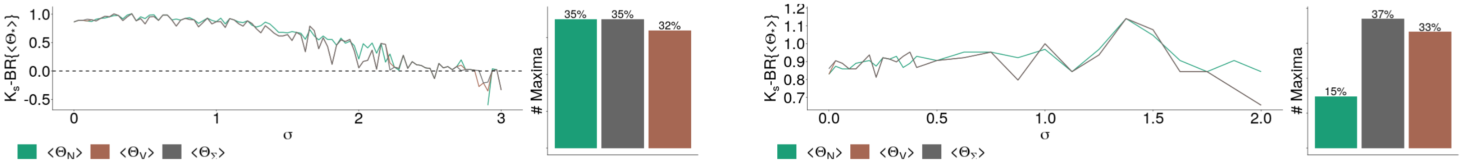


(d) Buishand range test.

Figure C.14: Trends in the lead times of the number of different opinion changes ($\langle \Theta_* \rangle$) under different change point tests. Left panels give results for disease infectivity $p = 0.2$, while panels on the right represent infectivity $p = 0.8$.

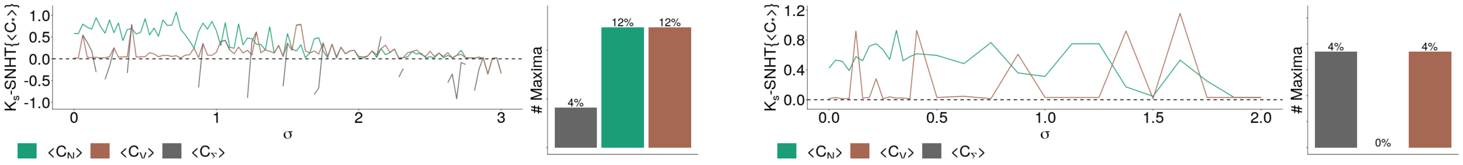


(a) Standard normal homogeneity test.

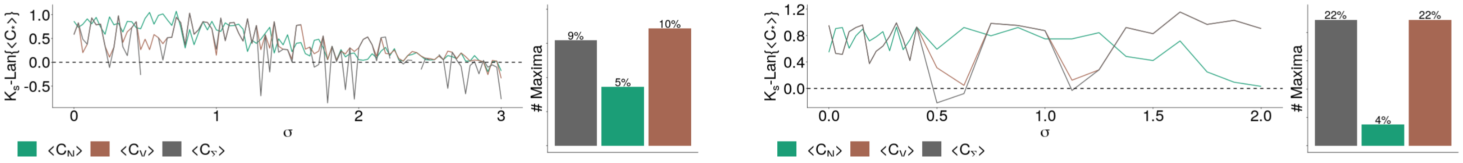


(b) Lanzante test.

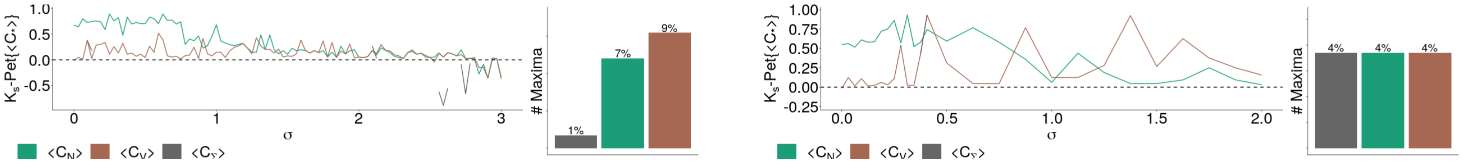


(c) Pettitt test.

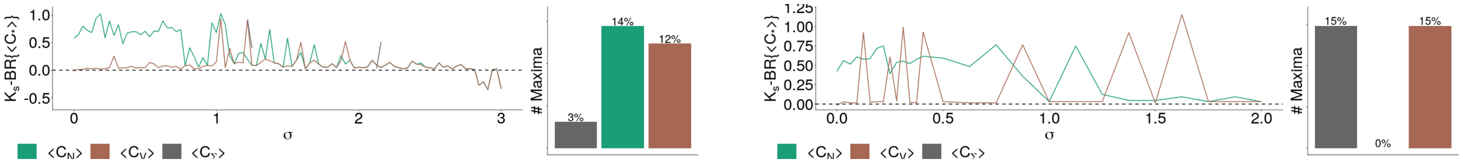


(d) Buishand range test.

Figure C.15: Trends in the lead times of the global clustering coefficient ($\langle C_* \rangle$) under different change point tests. Left panels give results for disease infectivity $p = 0.2$, while panels on the right represent infectivity $p = 0.8$.

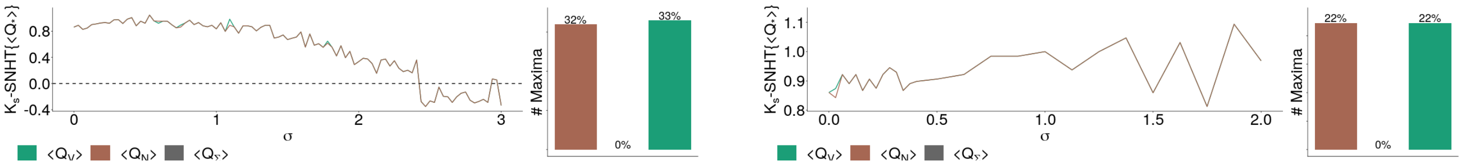


(a) Standard normal homogeneity test.

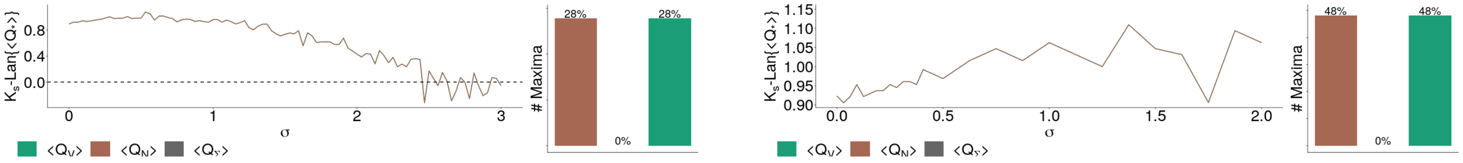


(b) Lanzante test.

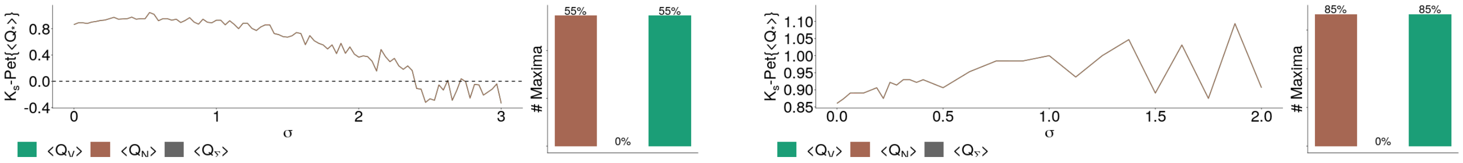


(c) Pettitt test.

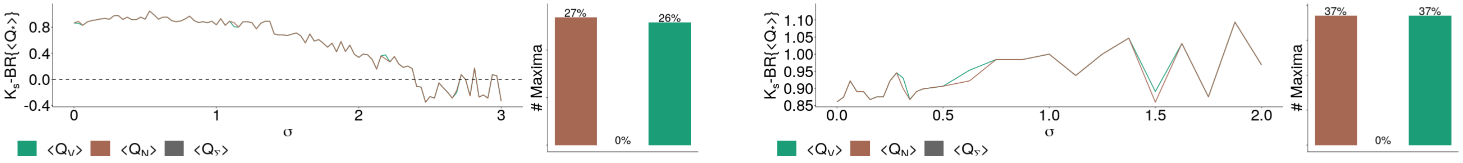


(d) Buishand range test.

Figure C.16: Trends in the lead times of the modularity statistic ($\langle Q_* \rangle$) under different change point tests. Left panels give results for disease infectivity $p = 0.2$, while panels on the right represent infectivity $p = 0.8$.

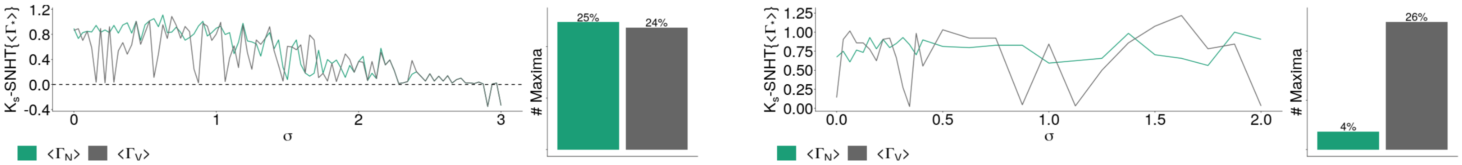


(a) Standard normal homogeneity test.

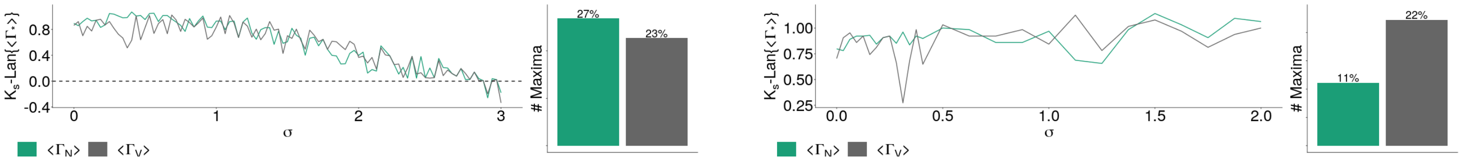


(b) Lanzante test.

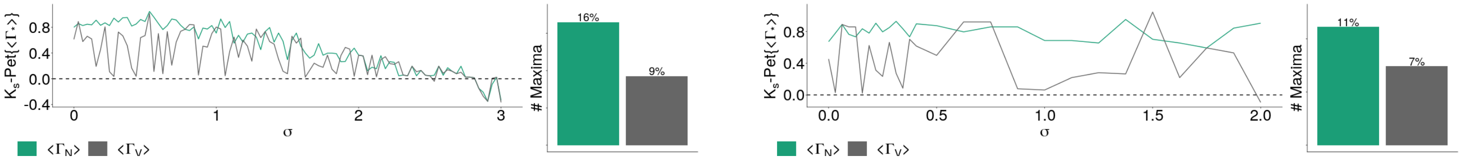


(c) Pettitt test.

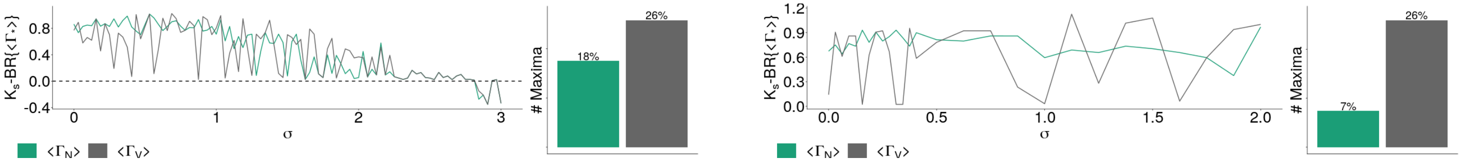


(d) Buishand range test.

Figure C.17: Trends in the lead time given by the probability of having an infected neighbour ($\langle \Gamma_* \rangle$) under different change point tests. Left panels give results for disease infectivity $p = 0.2$, while panels on the right represent infectivity $p = 0.8$.

#### Appendix D. EWS performance grid plots

All the figures in this section are grids showing the relative performance of each EWS with various change point detection tests and disease infectivities. Green tiles denote the social norm ($\sigma$) values for which the EWS gave the highest lead time, red tiles represent the smallest lead time, grey tiles represent lead times that are neither maxima nor minima and yellow tiles show where all EWS gave the same lead times. Black squares represent failed warnings (negative lead times) and white tiles represent undefined values (no lead time). In all figures in this section, panels (a) represents the infectivity $p = 0.2$ and panels (b) represent the infectivity $p = 0.8$.

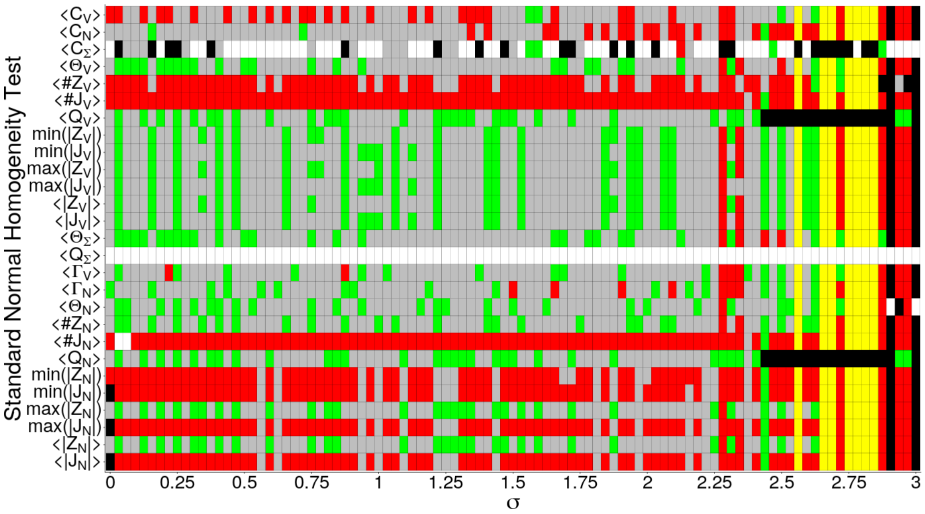


(a) Infectivity $p = 0.2$.

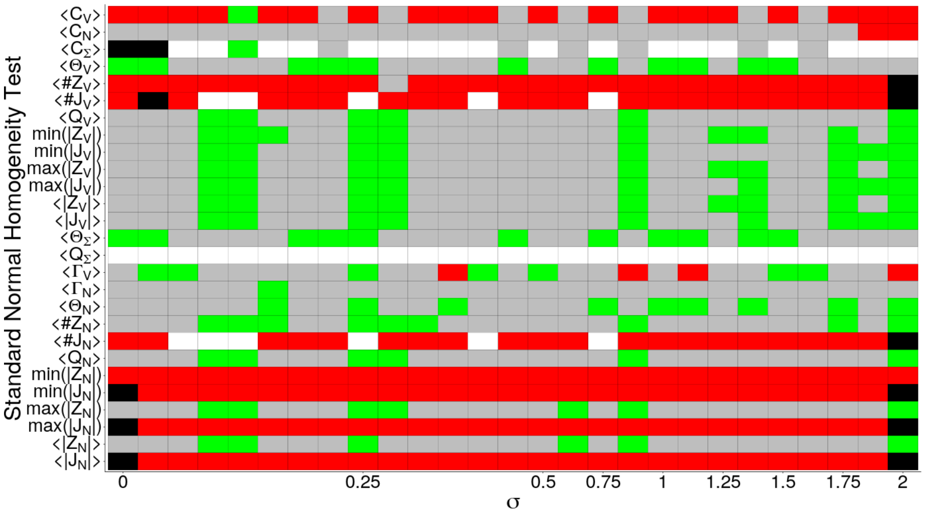


(b) Infectivity $p = 0.8$.

Figure D.18: Performance of each EWS per $\sigma$ value with the SNHT.

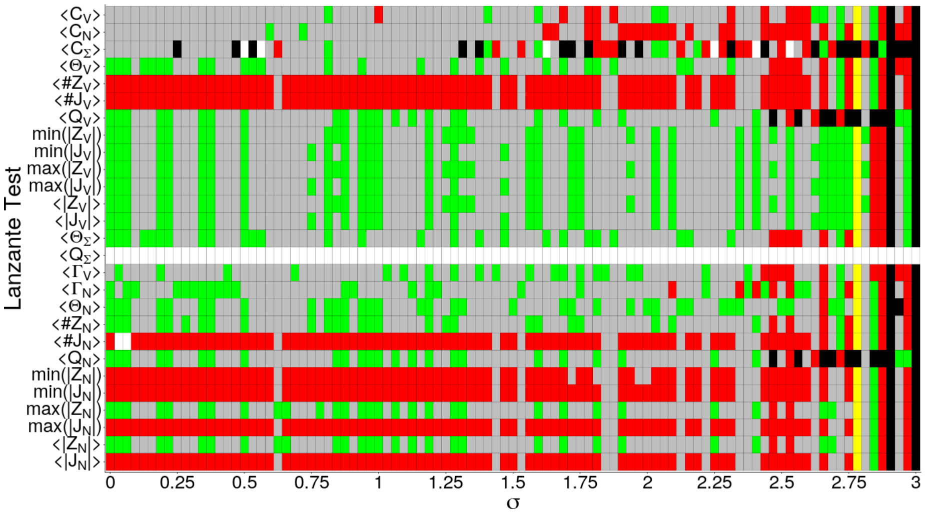


(a) Infectivity $p = 0.2$.

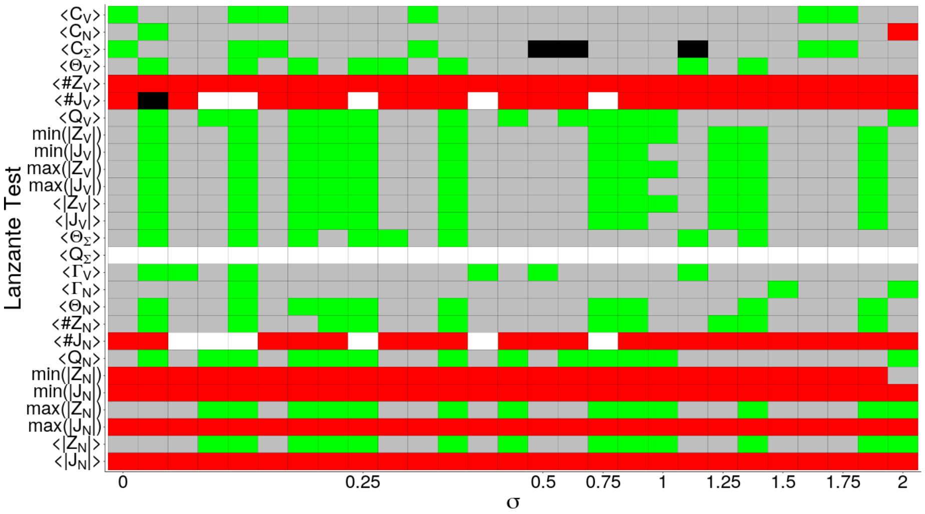


(b) Infectivity $p = 0.8$.

Figure D.19: Performance of each EWS per $\sigma$ value with the Lanzante test.

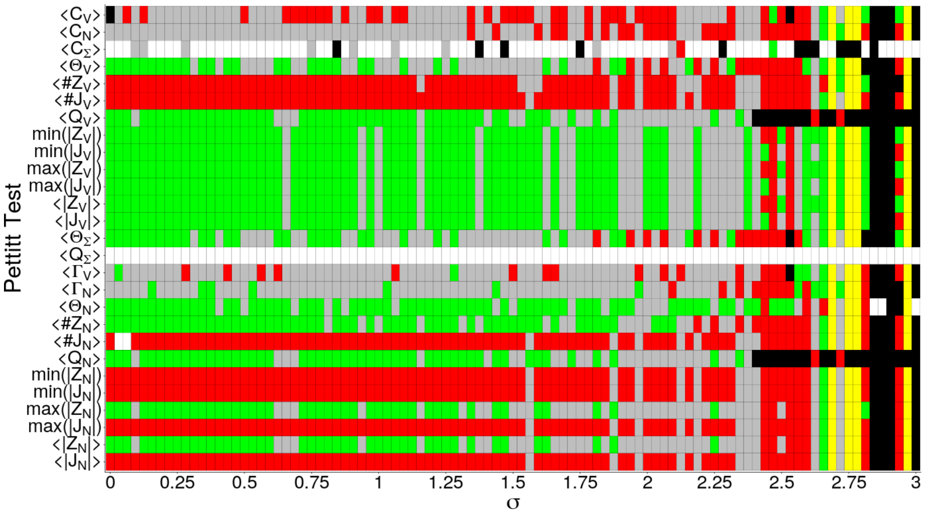


(a) Infectivity $p = 0.2$.

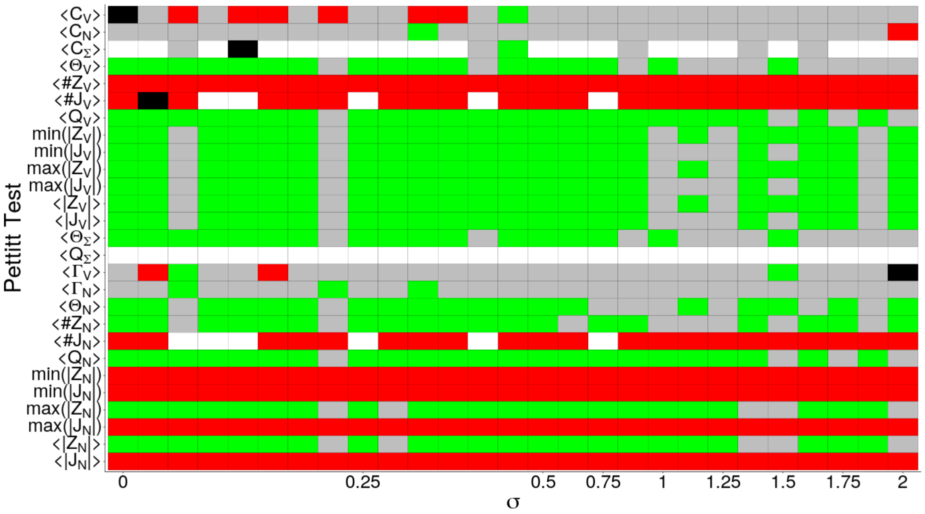


(b) Infectivity $p = 0.8$.

Figure D.20: Performance of each EWS per $\sigma$ value with the Pettitt test.

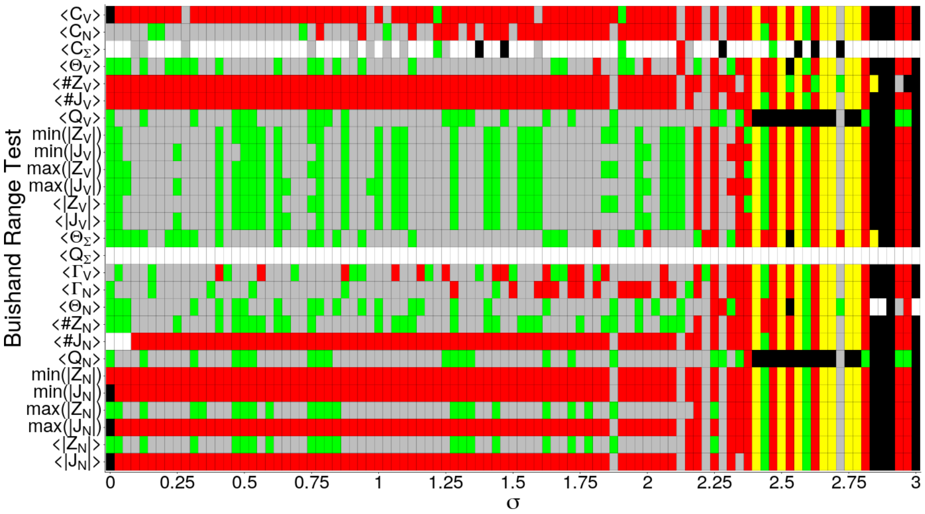


(a) Infectivity $p = 0.2$.

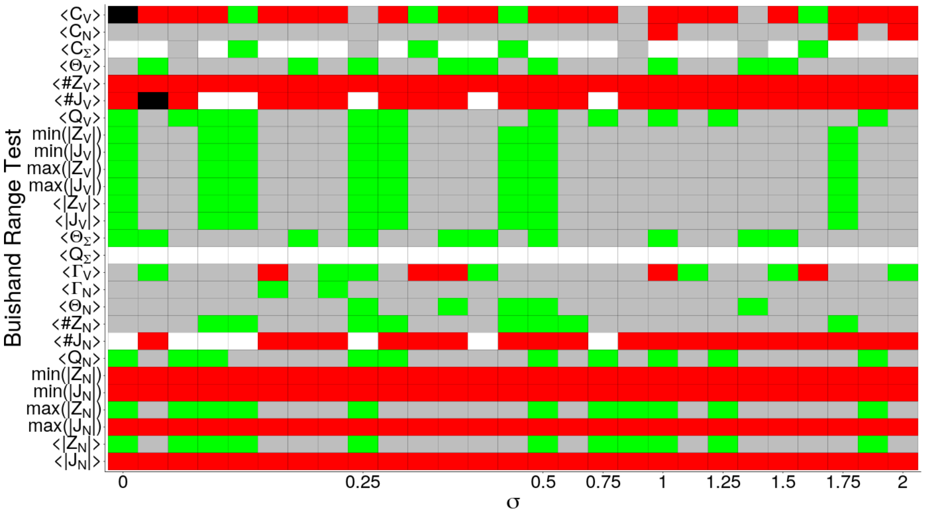


(b) Infectivity $p = 0.8$.

Figure D.21: Performance of each EWS per $\sigma$ value with the Buishand range test.

#### Appendix E. EWS performance bar charts

Grid showing the absolute and relative performance of each EWS. Green tiles denote the social norm $\sigma$ values for which the EWS gave the highest lead time, red tiles represent the smallest lead time, grey tiles represent lead times that are neither maxima nor minima and yellow tiles show where all EWS gave the same lead times. Black squares represent failed warnings (negative lead times) and white tiles represent undefined values (no lead time). In all figures in this section, panels (a) represents the infectivity $p = 0.2$ and panels (b) represent the infectivity $p = 0.8$.

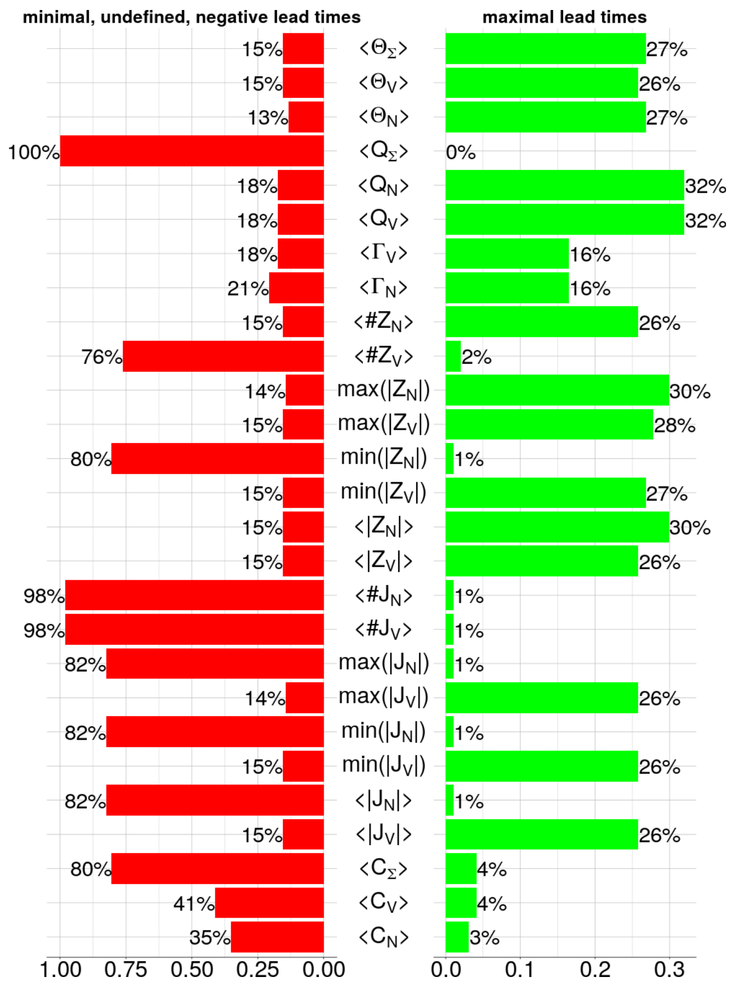


(a) Infectivity $p = 0.2$.

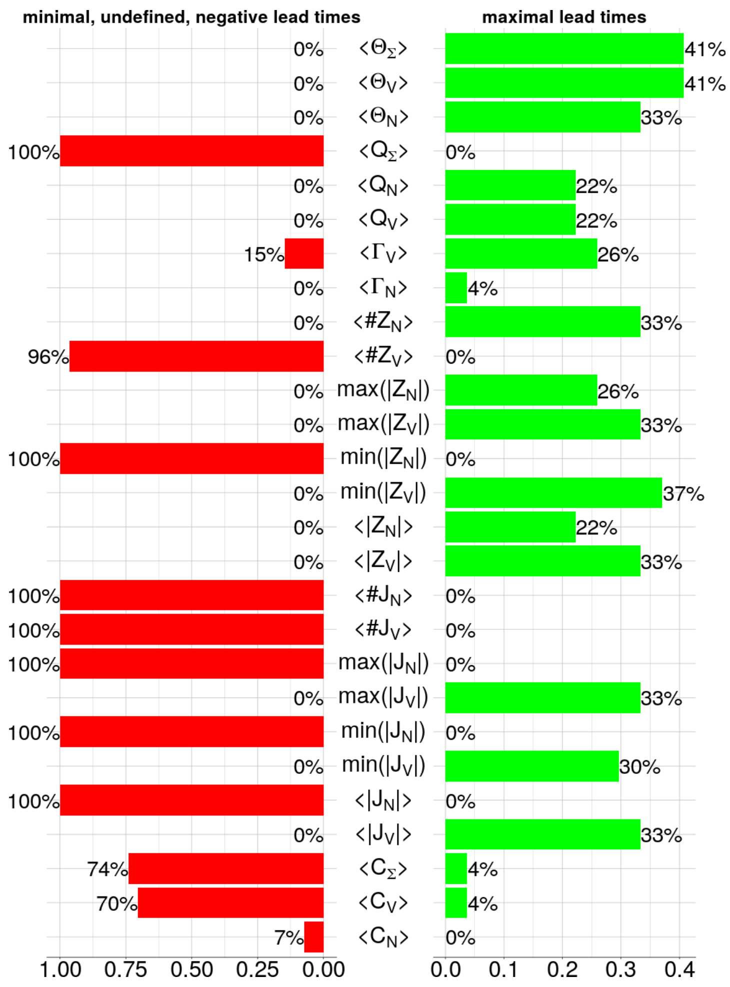


(b) Infectivity $p = 0.8$.

Figure E.22: Grids showing the absolute and relative performance of the EWS for different infectivities under the SNHT.

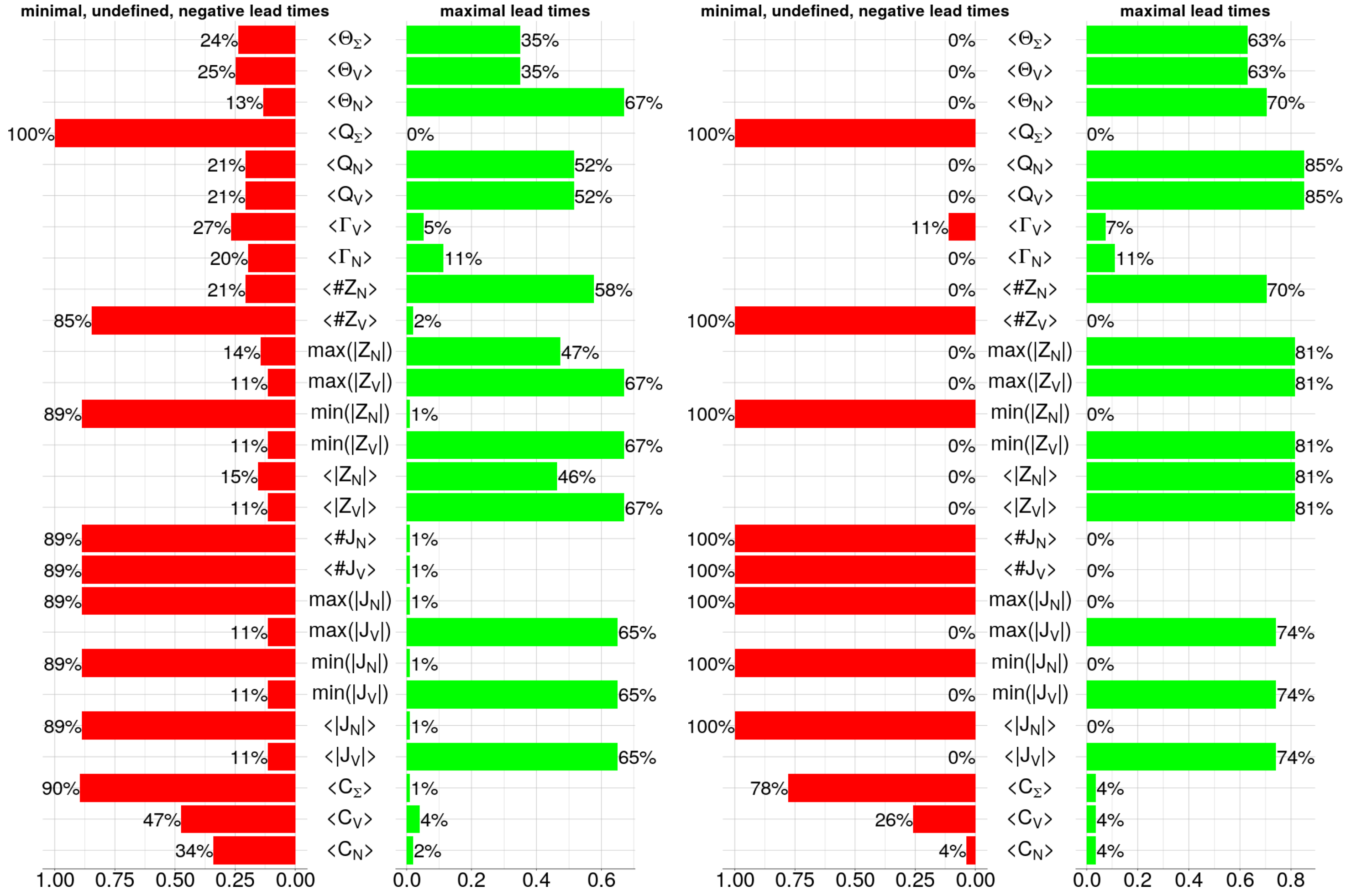


(a) Infectivity $p = 0.2$.

(b) Infectivity $p = 0.8$.

Figure E.23: Grids showing the absolute and relative performance of the EWS for different infectivities under the Pettitt test.

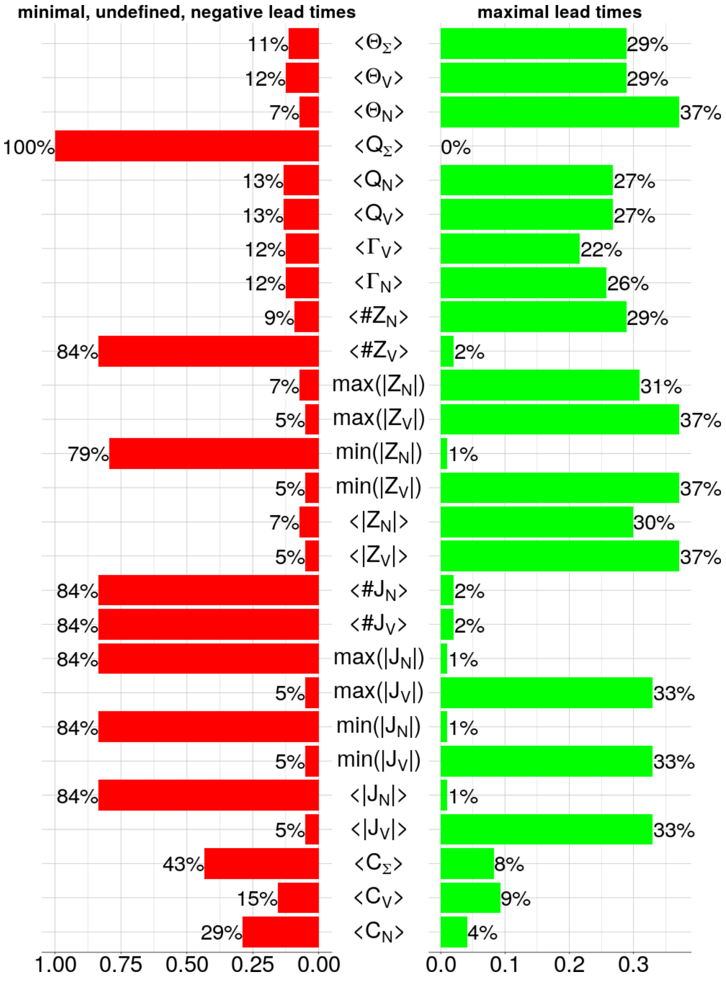


(a) Infectivity $p = 0.2$.

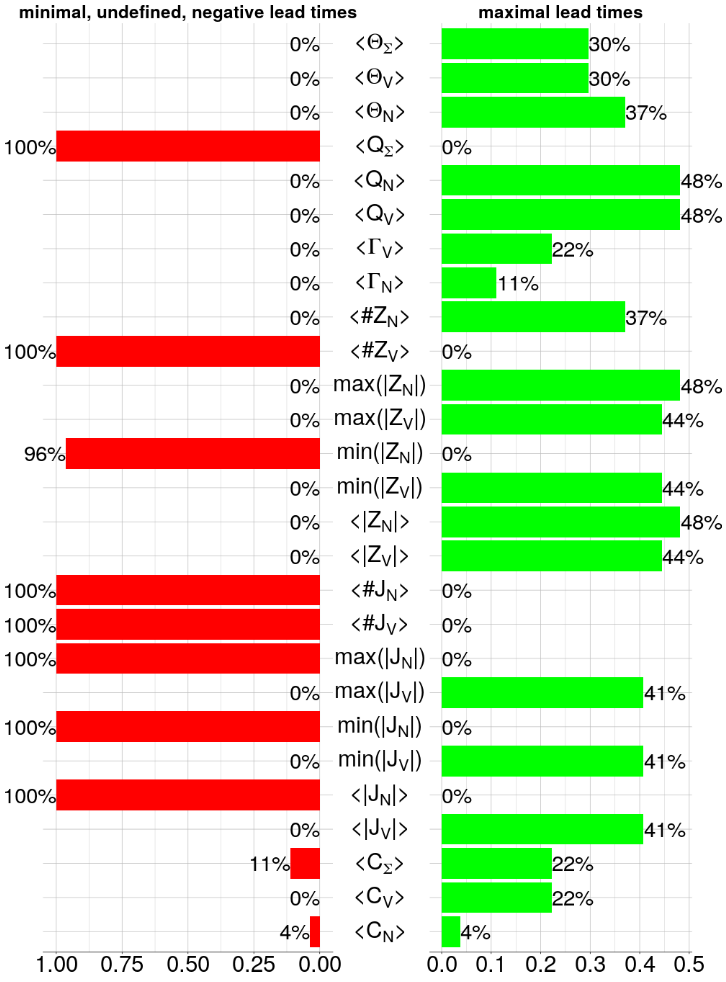


(b) Infectivity $p = 0.8$.

Figure E.24: Grids showing the absolute and relative performance of the EWS for different infectivities under the Lanzante test.

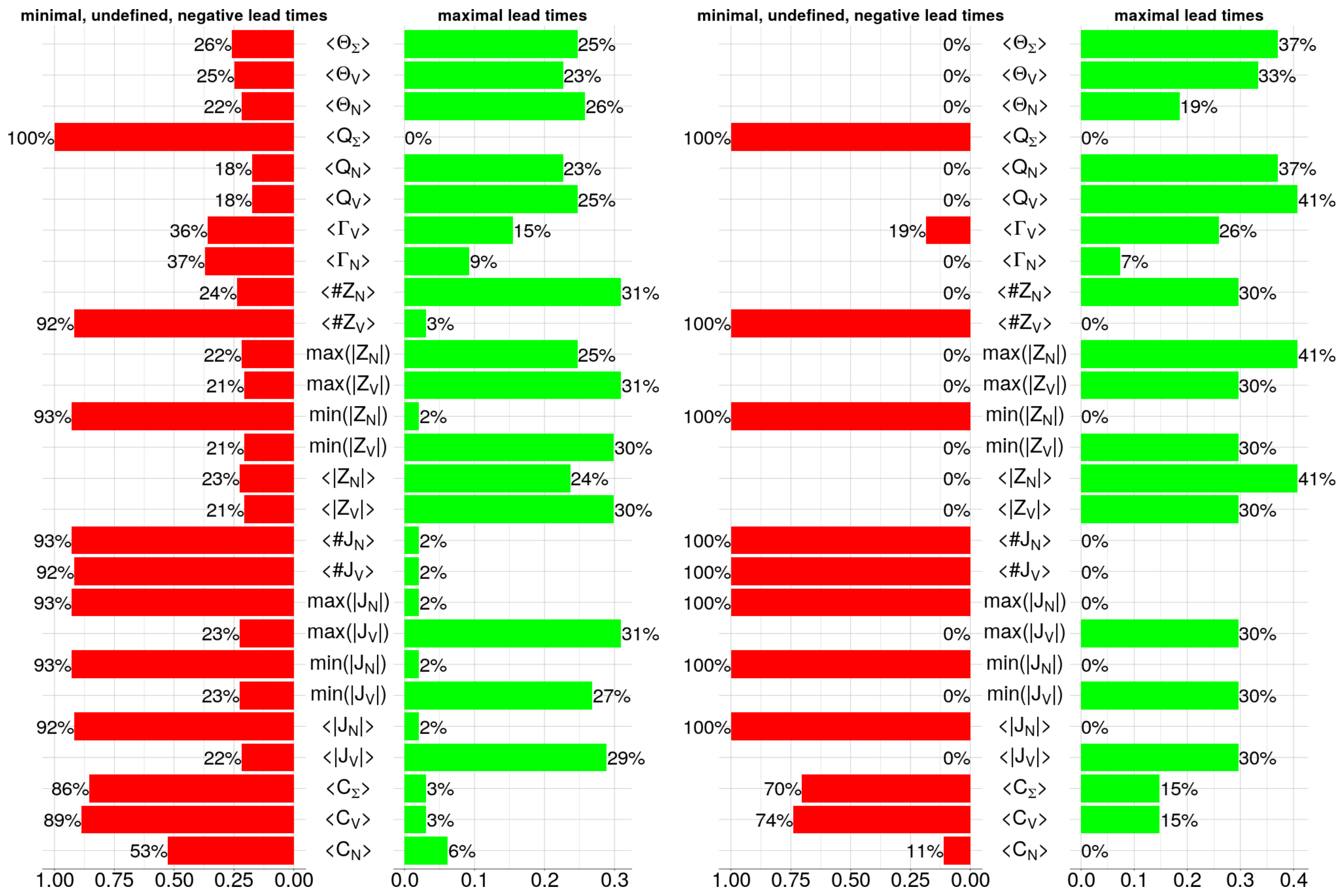


(a) Infectivity $p = 0.2$.

(b) Infectivity $p = 0.8$.

Figure E.25: Grids showing the absolute and relative performance of the EWS for different infectivities under the Buishand range test.
